# Evaluation of Contact-Tracing Policies Against the Spread of SARS-CoV-2 in Austria– An Agent-Based Simulation

**DOI:** 10.1101/2020.05.12.20098970

**Authors:** Martin Bicher, Claire Rippinger, Christoph Urach, Dominik Brunmeir, Uwe Siebert, Niki Popper

## Abstract

**Background:** Many countries have already gone through several infection waves and mostly managed to successfully stop the exponential spread of SARS-CoV-2 through bundles of restrictive measures. Still, the danger of further waves of infections is omnipresent and it is apparent that every containment policy must be carefully evaluated and possibly replaced by a different, less restrictive policy, before it can be lifted. Tracing of contacts and consequential breaking of infection chains is a promising strategy to help containing the disease, although its precise impact on the epidemic is unknown.

**Objective:** In this work we aim to quantify the impact of tracing on the containment of the disease and investigate the dynamic effects involved.

**Design:** We developed an agent-based model that validly depicts the spread of the disease and allows for exploratory analysis of containment policies. We apply this model to quantify the impact of divverent variants of contact tracing in Austria and to derive general conclusions on contract tracing.

**Results:** The study displays that strict tracing can supplement up to 5% reduction of infectivity and that household quarantine comes at the smallest price regarding preventively quarantined people.

**Limitations:** The results are limited by the validity of the modeling assumptions, model parameter estimates, and the quality of the parametrization data.

**Conclusions:** The study shows that tracing is indeed an efficient measure to keep case numbers low but comes at a high price if the disease is not well contained. Therefore, contact tracing must be executed strictly and adherence within the population must be held up to prevent uncontrolled outbreaks of the disease.

## 1 Introduction

Since in March 2020 the SARS-CoV-2 spread worldwide, countries managed to stop the exponential increase of case numbers several times already (for example, see situation report 88 by the World Health Organization (1)). Besides a few countries in Asia, such as China and South Korea, mostly countries in Europe have so far succeed in containing the disease due to swift and rigorous policy making of their governments. Hereby, they averted a potential overload of their health care systems, as happened in the Lombardy in March (2,3) or in Wuhan in January (4). Of these European countries including Germany, France, and Norway, Austria stands out by especially fast policy making in March which allowed the country to overcome the first disease wave very quickly.

Most of the introduced policies, such as closing schools, shops and restaurants proved to be effective in stopping the initial growth of the pandemic and led to a decrease in the new infections per day. However, due to social and economic reasons, lockdown policies cannot be upheld long enough to eradicate the disease completely (5). After a certain time, most of them must be lifted again, while other measures must be enforced to prevent a new upswing of the disease.

Testing and subsequent isolation of detected cases are the cornerstone of the disease containment program of COVID-19. Hereby, tests are used to evaluate the disease status of a suspected infected person, who is put under quarantine in case the test is found positive. The success of this strategy is directly coupled to the test sensitivity and the time it takes for a person to start to be infectious, get symptoms, initiate the test, make the test, and finally receive the test result – the less the better. Considering that this period usually spans a couple of days, secondary infections can be reduced but not entirely prevented this way.

The concept of contact tracing directly follows this idea. The essence of this strategy is to find and isolate those who might have already been infected by the newly confirmed case. Now, time is in favor of the regime since the tracer is looking for secondary transmissions and the disease is much less advanced in the affected persons. Consequently, it is now possible to find and isolate infected persons much earlier in their disease pathway, potentially even before they get infectious.

Although stigmatized as a violation of personal freedom, tracing is not always related to personal-data-tracking devices like mobile apps (6). Successful tracing of contacts starts by isolation of household members or by temporarily closing workplaces of confirmed SARS-CoV-2 infected persons. Many potentially infectious contacts can be traced by a simple interview with the patient as well.

Yet, besides many successfully detected and isolated newly infected persons, many entirely unharmed, healthy contact partners would also be put into quarantine this way which results in unintended adverse health effects and socioeconomic losses. This can be interpreted as drawback of the strategy.

Finding evidence that proves or quantifies the success of different tracing strategies is still difficult due to the novelty of the situation and simulation models currently being the only possibility to estimate the future impact of strategic changes. In Austria simulation results are systematically used by the ministry of health and public health authorities to guide health policy decision making and planning. The authors of this study are part of this process (7).

Hereby they apply an agent-based model (ABM) that is also subject of this study and fully documented in the Methods section and the Appendix. In contrast to typical aggregated compartment models such as the classic SIR model by Kermack and McKendrick (8), agent-based models (ABM) do not treat the populations as one continuously changing variable, but as the aggregate of individually modelled entities, so called agents (9,10).

Consequently, not only the transmission of the disease, but also policies like contact tracing can be modeled via agent-agent-interaction laws posing a very low level of abstraction compared to reality.

Key objective of this work is the qualitative and quantitative analysis of tracing as a containment policy for the COVID-19 pandemic. We ask which tracing strategy is the most successful, comes with the least socioeconomic costs, under which circumstances it works best, and if the disease can be contained by this measure alone or if we need additional policies.

## 2 Methods

For the development and analysis of our model, we followed the international guidelines of the ISPOR-SMDM Joint Modeling Good Research Practices Task Force (11,12) in the selection and justification of the model type as well as the description of methods and the reporting of our results.

We applied an agent-based modeling strategy in which each inhabitant is statistically represented by a model agent with certain demographic and disease related features. Disease transmission occurs via contacts between agents which occur inside of locations where agents interact with each other. The model is capable of introduction of certain policies, in particular different tracing strategies, that change the behavior of agents and/or transmission behavior of the disease.

### 2.1 Model Type

The chosen agent-based modeling type is a very complex approach for simulation of epidemics and there is a large variety of simpler simulation methods, such as the classic differential equation based SIR cohort model by Kermack and McKendrick (8). These strategies would be preferred if the sole purpose of the modeling process was the simulation of the disease. Yet, modeling of contact tracing policies requires modeling of person-to-person contacts which excludes population-based model types (see (12)). Consequently, a stochastic agent-based approach is necessary.

The model is comparable with similar models developed for Australia (13) and UK (14) but stands out by several features described in detail below. Briefly, (a) our model is based on a very accurate spatial and demographic image of the Austrian population in which each Austrian citizen is represented as an agent; (b) it utilizes a contact network based on different locations, such as households, workplaces and schools; and (c) it allows for tracing of agent-agent contacts and, consequently, for analysis of related tracing policies.

### 2.2 Decision-analytic Framework of the Policy Question

The target population of our study includes the entire population of Austria in the year 2020. The analytic time horizon of our analysis is February 21^st^ to November 15^th^, 2020, whereas the interval from February 21^st^ to April 9^th^ is used for calibration of the model and the time span between April 9^th^ and December 15^th^ is the actual simulation interval.

Although it has no direct application for the analysis of tracing measures, we will describe the initialization phase (February to April) of the model used for calibration as an additional model scenario as it reveals interesting insights into the epidemic.

For the actual evaluation of tracing policies, we investigate and compare 6 different strategies:1strategy without tracing (no-tracing), 3 strategies with location tracing (household tracing, workplace tracing, combined household and workplace tracing), and 2 different strategies of direct contact tracing. Since containment strategies for diseases are always a bundle of measures, we will evaluate each of the tracing strategies in combination with additional contact reduction. In this process, two different reduction values will be applied: one that will cause the case numbers to remain constant and one that will cause the numbers do decrease. For details, the reader is referred to Section 2.5.

We use our simulation to observe the timeline of the COVID-19 cases in different stages of the disease and treatment. We distinguish *pre-symptomatic* (infected within incubation period), *pre-confirmed* (sick persons waiting for a test / test result), *undetected* (infected which are and will never get tested due to no/mild symptoms), *confirmed* (cases confirmed by a positive test), as well as *quarantined* (cases put under quarantine due to a positive test) and *preventive quarantine* (persons put under preventive quarantine by a tracing policy). We track these numbers in three different forms: *active* (total number of persons in this state for a given point in time), *cumulative* (total number of persons that have ever been in this state since the start of the simulation) as well as *new* (number of persons that entered this state within the last day). To compare tracing strategies, we define the cost measure *quarantined per infection prevented* (*Qp*I*p*) which is calculated as

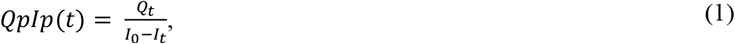

where I_0_ stands for the cumulative number (measured from the day of the policy introduction on May 15^th^) of *new infected* agents in the no-tracing scenario, I*t* describes the analogous number in the observed tracing strategy, and *Qt* stands for the cumulative number of *preventively quarantined* agents in the observed tracing scenario. This value can be interpreted as a cost function for the tracing strategy *t*.

The model uses a comparably high number of parameters which are identified using various data sources. Some values are determined using data from published literature, some are taken directly from census or routine data, some values are based on expert estimates, and some values require calibration routines.

Population parameters including fertility, mortality, and migration are identified via official publicly available census data from the Austrian national statistics office Statistics Austria. Parameters related to the contact behavior are primarily based on the POLYMOD contact survey (15), but open source data from Statistics Austria and the official data repository of the city of Vienna about household structure, employment rate, schools and workplaces have also been used (16–18).

COVID-19 related parameter values are based on recently published literature, expert opinions, the official disease reporting system of Austria and calibration processes. Time spans, such as incubation time, disease duration, etc., are parametrized using information from (19,20) and opinions from local virology experts. Hospitalization ratios and age distributions are gathered from statistical postprocessing of data from the official COVID-19 reporting system of Austria, the Epidemiologisches Meldesystem (EMS, (21)). The probability of an asymptomatic (undetected) disease progression is based on early antibody tests from Iceland (22). Finally, the infection probability and the impact of the already implemented lockdown measures in Austria were calibrated using the officially reported Austrian COVID-19 cases by EMS as a reference.

For more details on all parameters and all parameter values used the reader is referred to the Appendix, Section A1.3.3.

### 2.3 Model Specification

According to the Modeling Good Research Practices guidelines (11) we state a short, but easily understandable model description here. For a detailed and reproducible one, the reader is referred to the Appendix, Section A1.

The developed agent-based model is stochastic, population dynamic and it depicts every inhabitant of Austria as one model agent. It uses sampling methods to generate an initial agent population with statistically representative demographic properties and makes use of a partially event-based, partially time step (1 day) based update strategy to enhance in time.

#### Model Input

The model’s input consists of an event timeline of strategies that change the dynamics of the model at certain dates, mostly policies introduced by the government. In the simulation each element of the timeline is translated to one model-event that changes certain parameters or model mechanisms at the specified event-time. This input feature of the model makes it also possible to compare different strategies with each other, also regarding the introduction time of the strategy.

#### Model Initialization

Starting the simulation executes a population sampling routine, that has been implemented in the course of a prior research project (see DEXHELPP (23)). This sampling routine, a part of the Generic Population Concept (GEPOC, see (24)), ensures that the demography of Austria is well depicted by the agent-population, meaning statistically correct age, sex, and residence of each agent.

Moreover, the start date of the COVID-19 simulation model must lie between March 2020 and the current day and may be run arbitrarily long. To start the simulation at the defined point in time, say *t*_0_, the model applies an initialization simulation run from February 2020 until *t*_0_ to (a) calibrate the model parameters to the timeseries of the confirmed COVID-19 cases in Austria until *t*_0_, and (b) generate a valid initial agent-population for the actual simulation. This process guarantees that the correct number of susceptible, infectious, hospitalized, etc., model agents are present when the actual simulation is started.

The dynamics of the model are given by the interplay of four different submodels: a population, contact, disease, and policy model.

#### Population Model

First, agents are constantly affected by demographic change. Based on mortality and fertility rates, each agent has a certain time and age dependent probability to die or produce offspring any time during the simulation. Migration behavior is neglected in the model, due to very restricted immigration Austria in reality.

#### Contact Model

Moreover, the model makes use of a location specific contact model to establish interactions between agents. Agents meet daily at workplaces, schools, households and in leisure time and can transmit the virus with a certain transmission probability, if they are infectious.

#### Disease Model

After being infected, agents go through a detailed disease and/or patient pathway that depicts the different states of the disease and the treatment of the patient. This pathway is modeled via specifically distributed transition times and is not Markovian. It contains branches with respect to disease severity (asymptomatic or symptomatic, etc.), and with respect to treatment (hospitalization or home isolation, normal bed or intensive care unit, etc.) and always ends in the recovery or death of the agent. The disease and treatment states influence the contact behavior of the agent and, consequently, how it can transmit the virus.

#### Policy Model

Clearly, not only the disease and treatment state, but also the currently active policies influence the contact behavior of agents. Policies may lead to closure of certain locations, which make them unavailable for contacts, but may also cause a reduction of contacts or a reduction of the infectivity of contacts due to increased hygiene. Focus of the present study are tracing policies which cause additional home-quarantine for contacts of newly infected agents, which removes them from the contact network and reduces their capabilities to transmit the disease.

#### Model Output

The outcomes of the model are time series with a daily time basis. They consist of aggregated numbers describing the current nation- and/or region-wide spread of the disease as well as numbers depicting the contact behavior of agents. These include, for example, the cumulative number of confirmed cases, the number of currently active asymptomatic cases, the total number of daily newly infected 10- to 30-year-old females, the total number of daily contacts for school children, or the average number of secondary infections per agent (=*R*_*eff*_). The outcomes observed for this study in particular are explained in Section 2.2.

### 2.4 Model Implementation

The simulation of ABMs like the specified agent-based COVID-19 model is a huge challenge with respect to computational performance. As the model cannot be scaled down, almost 9 million interacting agents need to be included into the model to simulate the spread of the disease in the entire population of Austria.

These high demands exclude most of the available libraries and software for agent-based modeling including AnyLogic, NetLogo, MESA, JADE, or Repast Simphony (25–29). Most of these simulators cannot be used as their generic features for creating live visual output generates too many overheads.

Consequently, we decided to use our own agent-based simulation environment ABT (Agent-Based Template, see (30)), developed in 2019 by dwh GmbH in cooperation with TU Wien. The environment is implemented in JAVA and specifically designed for supporting reproducible simulation of large-scale agent-based systems. More technical details are found in the Appendix, Section A3.

### 2.5 Strategies and Scenarios

In this section, we describe the tracing strategies and compliance scenarios mentioned in Section 2.2 and define them in the upcoming four sections: The initialization phase of the model is defined in section 2.5.1, the reference strategy without tracing and the three strategies dealing with location tracing are defined in Section 2.5.2, and two strategies with different types of individual tracing are specified in Section 2.5.3. The concept for additional contact reduction policies is presented in Section 2.5.4.

At some points, the specification of the strategies and scenarios are not fully reproducible to support readability. For tables containing the precise, reproducible model parametrization we refer to the Appendix, Section A4.

#### 2.5.1 Definition of the Initialization Phase

We chose April 9^th^, 2020, 08:00 AM as the initial time of our actual simulation – we will henceforth denote this time as *t*_0_. To start the simulation at this date, it is necessary to run an initialization phase that validly depicts the entire progression of the disease until this date and save the final state of this phase as an input to the actual simulation. Interestingly, this initialization phase also reveals certain features about the disease which cannot be measured in the real system, for example the time series of the asymptomatic cases. Consequently, we decided to include this initialization phase to this study as initial scenario.

By April 9^th^, countrywide lockdown in Austria had already managed to reduce *R*_*eff*_, the effective transmission rate of the disease, below 1, causing the number of newly infected people per day to decrease. About 12,900 positive virus tests had been reported until this date ^1^. To guarantee that the final state of the initialization phase matches this number, a calibration process was performed adjusting both infectivity and impact of lockdown policies. This process is, in more detail, described in the Appendix, Section A1.3.4. The country implemented nationwide closure of schools and workplaces on March 16^th^, yet our calibration process revealed that this lockdown should rather be modelled as a process with several steps, which are briefly listed in the Appendix, Table A7. It is apparent, that the modelled policy events and, in particular, their parametrization cannot be taken into account separately – some of them might have a larger, some a smaller impact in reality than in the model – yet the summary of all policies allowed us to calibrate the current curve of the disease by feasible and causally founded assumptions.

#### 2.5.2 Definition of No-Tracing and Location Tracing Strategies

In the simulation, all tracing policies have been implemented on May 15th, a time at which the new upswing of the epidemic can already be observed by an increasing number of new infections, independent of the compliance level.

To create a reference for evaluation of tracing strategies, we specified a no-tracing strategy in which no tracing is present whatsoever. As soon as infected person agents become confirmed cases, they isolate themselves, but there are no consequences for contact partners whatsoever.

As the first public health measure to evaluate we established so-called location tracing policies. We define this policy as the reaction of a person’s direct surrounding in response to a positive SARS-CoV-2 test result. While isolation of the affected person is done as usual, now also all persons in the direct surrounding of the infected person will become isolated as well, independent of their current disease state. In this process, the surrounding is defined as the group of persons that commonly visit the same locations as the infected person. By this measure we expect to find and isolate a high percentage of infected persons before they even become visible to the system.

In the model, we studied the effects of location tracing regarding two location types: household and workplace. The policy *household tracing* means that as soon as an agent enters the confirmed status, all other members of the agent’s household are isolated as well. In *workplace tracing*, the workplace of a confirmed COVID-19 patient is temporarily closed, and all the coworkers are put into preventive quarantine.

In isolation, agents only have contacts with other members of their household. They do not attend school or work and do not have leisure time contacts. After a fixed number of days – we chose 14 days for our strategies – agents are released from isolation and can resume their normal behavior, if they turn out to be unaffected by the virus. Clearly, the availability of a precise test could reduce the required quarantine length, yet this feature is not included in the model thus providing conservative estimates.

We evaluated the impact of the location tracing for households and for workplaces separately as well as in combination, henceforth denoted as *combined tracing* strategy.

#### 2.5.3 Definition of Individual Tracing Strategies

Extending the ideas of location tracing, we studied the effects of *individual tracing* of contacts. For this tracing policy, we assume that a certain amount of people records their contacts outside of their household, for instance, by using a tracing app on their smartphone or on a similar device. In this process, a contact is recorded if both involved persons use the tracing device. We assume that the tracing is completely accurate. In this way, all contacts between persons using the tracing device are recorded and, most importantly, there is no infection between two tracing people that goes undetected. These contacts are saved for a specific recording period. If a person using the tracing device becomes a confirmed case of COVID-19, the recorded contacts are informed and placed under preventive quarantine. The implications of the preventive quarantine are the same as in Section 2.5.2.

The effectiveness of this policy has been evaluated on top of the location tracing policies for households and workplace contacts, that is, the *combined tracing* strategy. We considered rates of 50% and 75% of people using the tracing device and a recording period of 7 days. The length of the preventive quarantine is fixed at 14 days.

#### 2.5.4 Definition of Stagnation Levels and Contact Reduction Strategies

When evaluating the impact of a containment policy via modeling and simulation two difficult problems emerge:

First, the policy lacks a comparator. It is not fair to assume that governments will not constitute alternative measures if the policy is not found suitable. Hence, a comparison between a policy and the correspondent no-policy strategy, as typically done in (health) technology assessment studies, is not correct.

Second, the policy cannot be evaluated without thinking about additional measures that are active at the same time. Consequently, scenarios with assumptions for additional policies need to be made to evaluate the impact of the policy in the context of other measures. This might also cause problems since not every policy is suitable in combination with any other policy or for any state of the pandemic. In particular, the analyzed contact tracing policies are not applicable if the case numbers exceed a certain boundary due to limited human resources.

To overcome these two problems, we define the *stagnation level* of a tracing policy by the percentage of additional reduction of disease infectiveness in leisure-time, workplaces, and schools necessary to keep the disease numbers on a constant level, that is, they neither decrease nor increase. The higher the stagnation level, the more additional policies need to be introduced for disease containment, and the less effective is the original policy.

Technically, finding these stagnation levels is related to a calibration process. In this process, the calibration value is the parameter of an *infectivity reduction event* scheduled for leisure time, workplace, and schools which is scheduled for May 15^th^, that is, the same date as the introduction of the tracing policy. The value is calibrated between 100% (contacts are not infectious anymore) and 0% (contacts are fully infectious, that is, as infectious as calibrated in the initialization phase scenario). A standard bisection method is used with a Monte Carlo simulation (12 runs each) in the loop. Since stagnation of the case numbers is the goal of the calibration process, the target value of the calibration routine is defined as the slope of the regression line fitted through the simulated timeseries of the *new confirmed cases per day* between June 1^st^ and July 1^st^.

Figure 1 gives an image of the calibration process for the stagnation levels of the no-tracing policy. The case numbers for the time between April 9^th^ and May 1^st^ are dropping due to the upheld lockdown measures. On May 1^st^, all containment measures have been lifted and the case numbers are rising again. On May 15^th^, the *infectivity reduction event* damps this upswing and is calibrated to cause stagnating numbers. The calibration routine terminates for 74.58% infectivity reduction in leisure time, workplaces and schools which causes the case numbers to stagnate at about 390 *new confirmed cases per day*. According to the specified ratio of undetected cases this corresponds to about 2000 *new infected cases* per day, as depicted in Figure 2.

**Figure 1.**
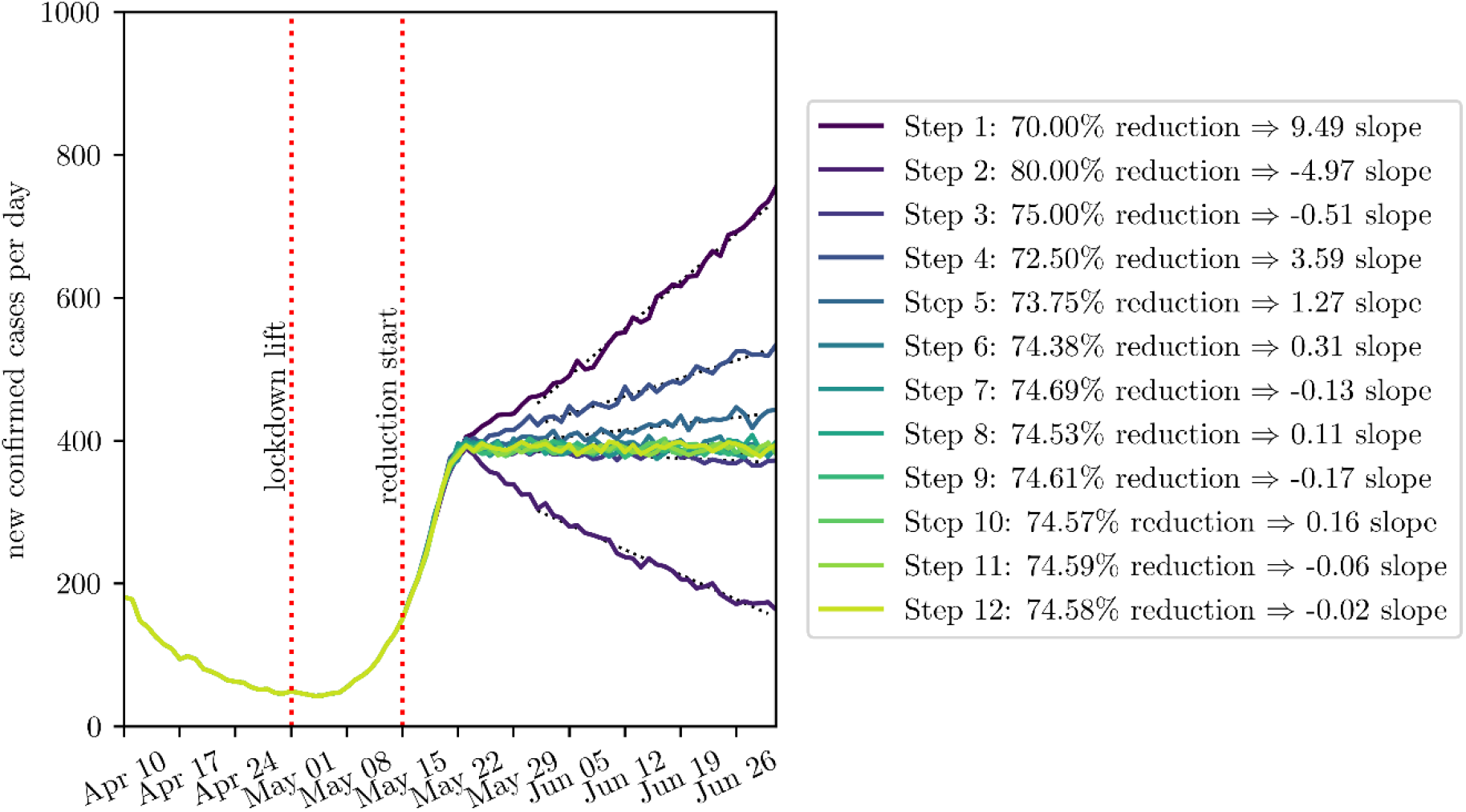
Calibration process of the stagnation level for the *no-tracing* strategy. The colormap indicates the sequence of bisection steps the calibration routine has performed with respect to the varied parameter: the infectivity reduction on May 15^th^. The black dotted curves show the regression lines used for evaluating the calibration target: the slope of the regression line.

**Figure 2.**
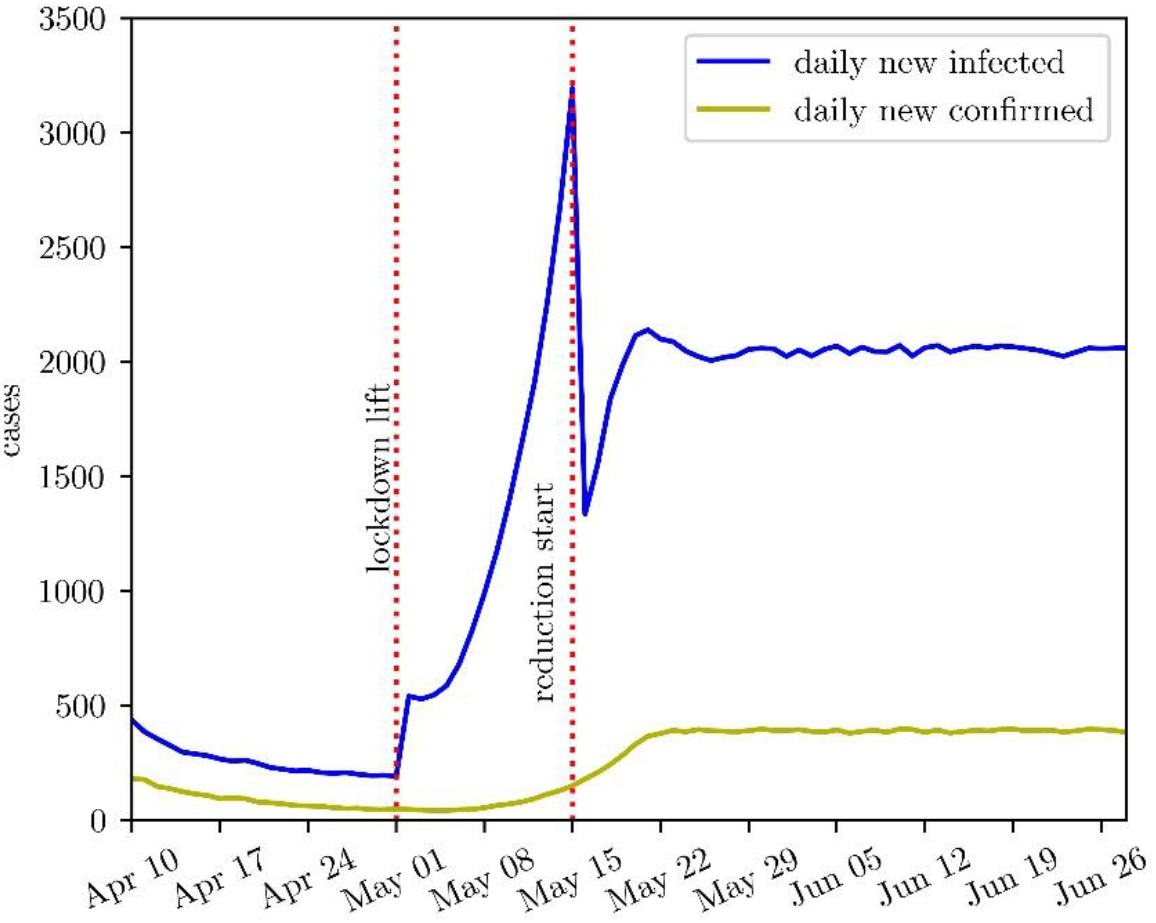
Curves for the *daily new confirmed* and the *new infected* cases for the *no-tracing* strategy when calibrated to the stagnation level (74.58%). The lift of the lockdown as well as the introduced infectivity-reduction instantaneously reduces the level of *new infections* while the change for the *new confirmed* cases happens more smoothly and time delayed.

Note that the bisection algorithm does not always generate a fully monotonically converging result due to the stochasticity of the simulation. Even though Monte Carlo simulation is used to filter the effects of the stochastic model, a small level of perturbation remains. Consequently, the results of the algorithm are precise up to the second decimal.

In the following we use the concept of stagnation levels to compare the tracing policies. First, we determine the stagnation levels of all six tracing policies and compare these with each other. This way, we will find out how much contact reduction can be compensated by which tracing policy. In a second step, we will simulate each of the six tracing policies together with the stagnation level of the no-tracing strategy (depicted above). This way, we will determine how different tracing strategies affect the decline of the case numbers<u>2.5.</u>

## 3 Results

### 3.1 Initialization Phase

On the average, by March 16^th^, the modelled total contacts per day were reduced by about 78%, with additionally reduced infectivity of contacts at workplace and in leisure time by 50%. That is, the march lockdown in Austria resulted in a total infectivity reduction of the disease by about 89% (=1− (1− 0.78)(1 − 0.5)).

Figure 3 depicts the results of the initialization phase. It nicely displays that the *confirmed* cases are only one part of the total infected population and that it is necessary to consider all of them to generate a feasible initial population at *t*_0_: the *undetected*, that experience no or only mild symptoms and are not confirmed by a test, the *pre-symptomatic*, that are still within the incubation period, and the *pre-confirmed*, that wait for being tested or have not yet received their positive test result.

**Figure 3.**
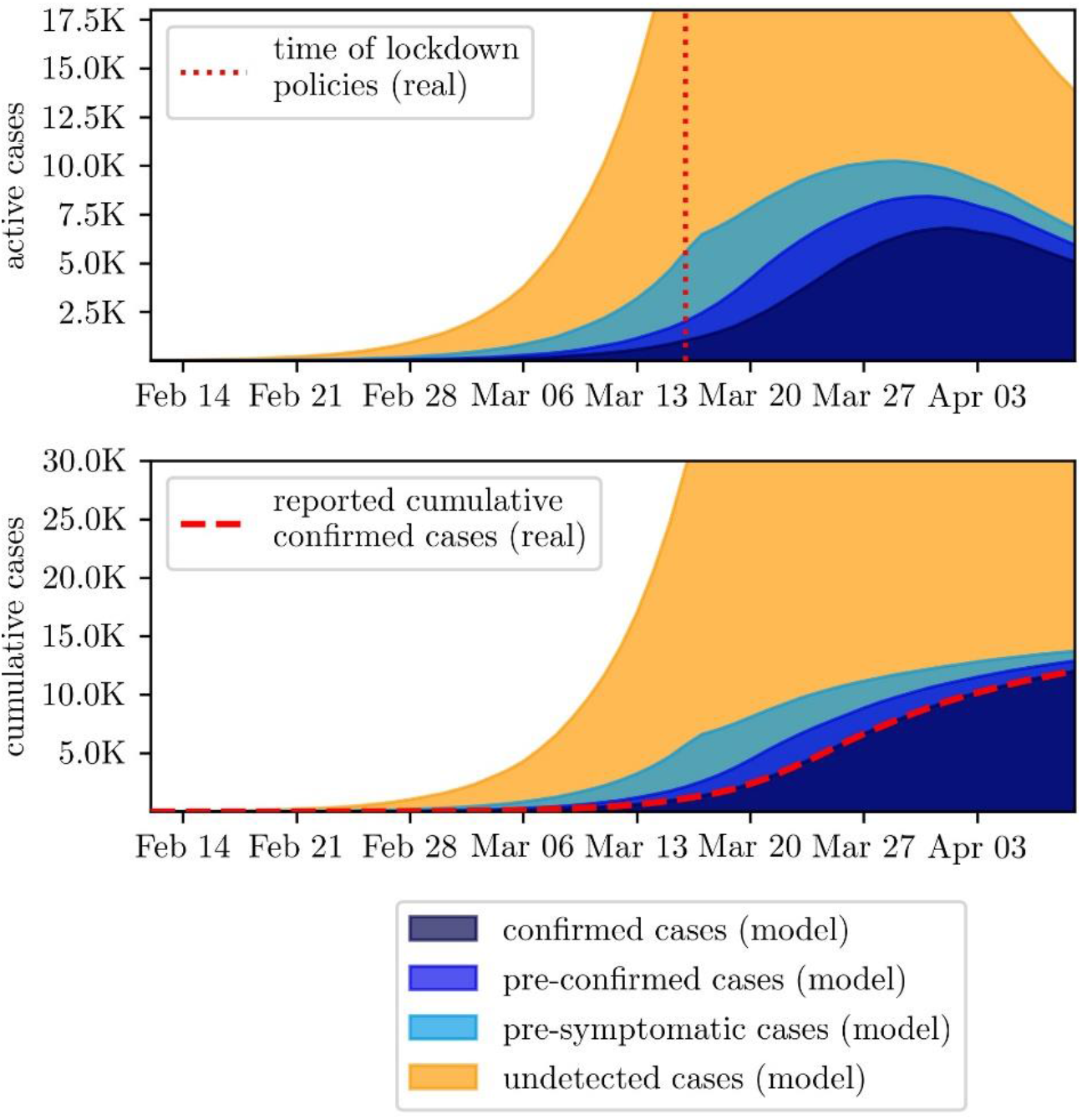
Comparison of the initial phase and reported data from Austria. Upper plot shows active cases while the lower plot displays cumulative.

For calibration purposes a bisection algorithm was applied to iteratively improve the value of one parameter value after the other. This strategy is possible as the impact of the calibrated parameters can be measured at different points in time: the base *infection probability* can be calibrated in the period before introduction of measures, the impact of the first policy can be calibrated in the period between the first and the second, etc. Hence the multi-dimensional calibration problem can be decoupled into several scalar ones. More on this strategy is found in the Appendix, Section A1.3.4.

### 3.2 Stagnating Case Numbers

In the first comparison of tracing policies, we used the concept of stagnation levels introduced in Section 2.5.4. We used the presented calibration routine for all six strategies and determined the corresponding stagnation levels. They are summarized Table 1

**Table 1.** Calibrated stagnation levels for all six contact tracing strategies. Higher stagnation levels indicate that more contact/infectivity reduction policies need to be added to the tracing strategy to contain the disease.

| Tracing strategy | Stagnation level (%) | Stagnation level improvement compared to no-tracing (%) |
| --- | --- | --- |
| *no-tracing* | 74.58% | - |
| *household tracing* | 72.96% | 1.62% |
| *workplace tracing* | 73.53% | 1.05% |
| *combined tracing* | 71.47% | 3.11% |
| *50% individual tracing* | 70.46% | 4.12% |
| *75% individual tracing* | 69.40% | 5.18% |

Moreover, we compared the simulation results for all six tracing strategies together with the correspondent infectivity reduction on stagnation level. While the curves of the *new confirmed* cases all show the same picture as the one seen in Figure 2, the numbers of *new preventively quarantined* agents differ due to different strictness of the tracing policies. This is displayed in Figure 4. The scenarios involving workplace tracing show an unsteady increase in quarantined agents directly after start of the policy. This results from instantaneously putting all workplaces containing confirmed infected agents under quarantine the moment the policy is introduced. Analogous to the *new confirmed* cases, the *new quarantined* agents also converge to equilibrium values which correspond to the strictness of the tracing strategy.

**Figure 4.**
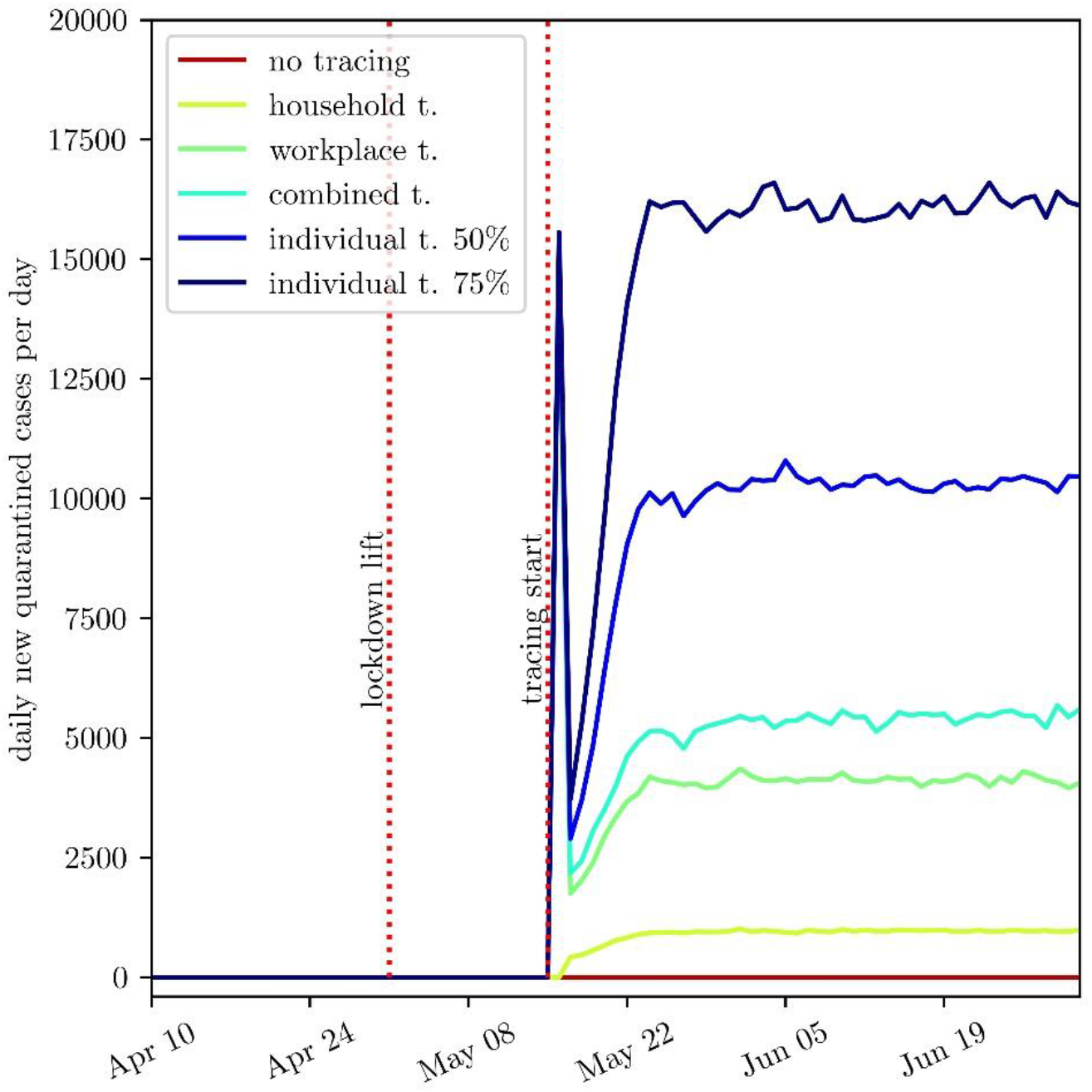
Simulation results for the new preventively quarantined agents, that is agents put under preventive quarantine as a traced contact. All six tracing strategies are evaluated in combination with the corresponding infectivity reduction for reaching the stagnation level

### 3.3 Decreasing Case Numbers

In a second step we evaluate the performance of the tracing strategies for a contained scenario. To do this, we fix the infectivity reduction on the stagnation level of the no-tracing strategy, that is 74.58%, and run the simulation for each of the 6 tracing strategies. The curves for the *new confirmed* cases are displayed in Figure 5. Clearly result curves start to diverge from May 15^th^ since policies are introduced at this date. Thereafter, more strict tracing policies cause a faster decline of the case numbers. Results including information about the preventive quarantined persons are summarized in Table 2, including the cost measure *QpIp* defined in (1).

**Table 2.** Summary of simulation results for all six tracing strategies simulated together with 74.58% infectivity reduction.

| Tracing strategy | *cumulative infected agents*<br>(May 15<sup>th</sup> - Nov 15th) | difference to<br>*no-tracing*<br>strategy | *cumulative preventively quarantined agents* (May 15<sup>th</sup> - Nov 15th) | *quarantined per infection prevented*<br>($QpIp$) |
| --- | --- | --- | --- | --- |
| *no-tracing* | 371 709 | - | - | - |
| *household tracing* | 220 046 | 151 663 | 104 182 | 0.69 |
| *workplace tracing* | 244 092 | 127 616 | 530 078 | 4.15 |
| *combined tracing* | 165 167 | 206 542 | 458 006 | 2.22 |
| *50% individual tracing* | 141 822 | 229 887 | 735 792 | 3.2 |
| *75% individual tracing* | 119 826 | 251 883 | 970 461 | 3.85 |

**Figure 5.**
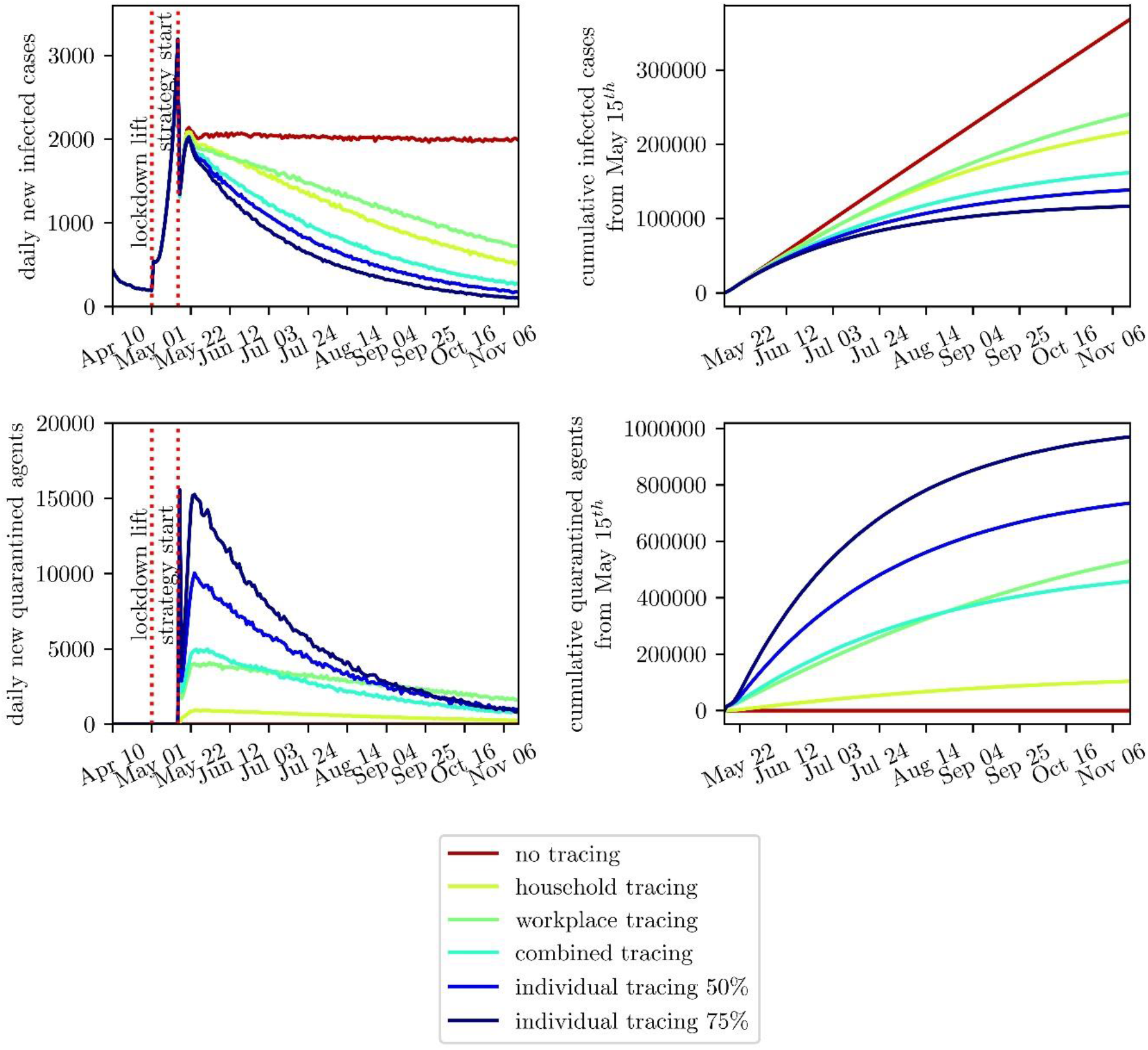
Simulation results for all six tracing strategies simulated together with 74.58% infectivity reduction are displayed. The left plots show the *new confirmed* and *new preventive* quarantined cases. The right plots display the correspondent cumulative curves, calculated from May 15^th^.

Figures 6 and 7 give an image of the cost measure showing the cumulative quarantined and the prevented infections in relation to each other. In specific, Figure 7 clearly displays that the ratio between these values is time dependent. The corresponding values are found in Table 3.

**Table 3.**
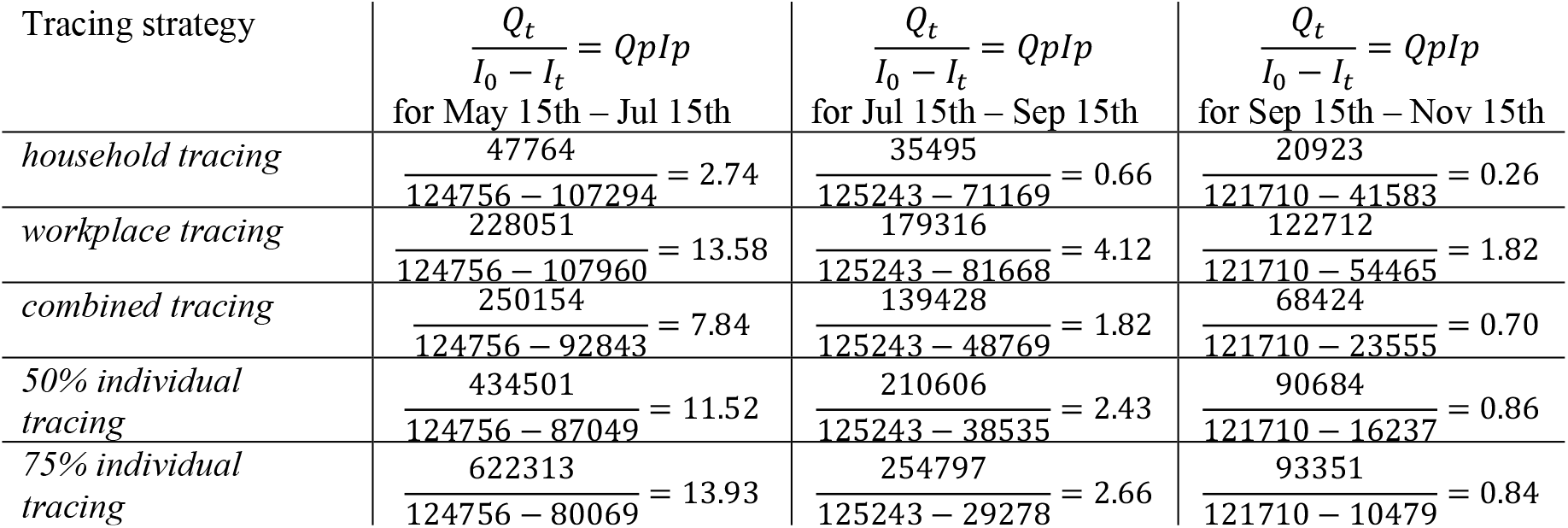
Change of the *quarantined per infection prevented* (QpIp) value over time for the different strategies

| Tracing strategy | $\frac{Q_t}{I_0 - I_t} = QpIp$<br>for May 15th – Jul 15th | $\frac{Q_t}{I_0 - I_t} = QpIp$<br>for Jul 15th – Sep 15th | $\frac{Q_t}{I_0 - I_t} = QpIp$<br>for Sep 15th – Nov 15th |
| --- | --- | --- | --- |
| household tracing | $\frac{47764}{124756 - 107294} = 2.74$ | $\frac{35495}{125243 - 71169} = 0.66$ | $\frac{20923}{121710 - 41583} = 0.26$ |
| workplace tracing | $\frac{228051}{124756 - 107960} = 13.58$ | $\frac{179316}{125243 - 81668} = 4.12$ | $\frac{122712}{121710 - 54465} = 1.82$ |
| combined tracing | $\frac{250154}{124756 - 92843} = 7.84$ | $\frac{139428}{125243 - 48769} = 1.82$ | $\frac{68424}{121710 - 23555} = 0.70$ |
| 50% individual tracing | $\frac{434501}{124756 - 87049} = 11.52$ | $\frac{210606}{125243 - 38535} = 2.43$ | $\frac{90684}{121710 - 16237} = 0.86$ |
| 75% individual tracing | $\frac{622313}{124756 - 80069} = 13.93$ | $\frac{254797}{125243 - 29278} = 2.66$ | $\frac{93351}{121710 - 10479} = 0.84$ |

**Figure 6:**
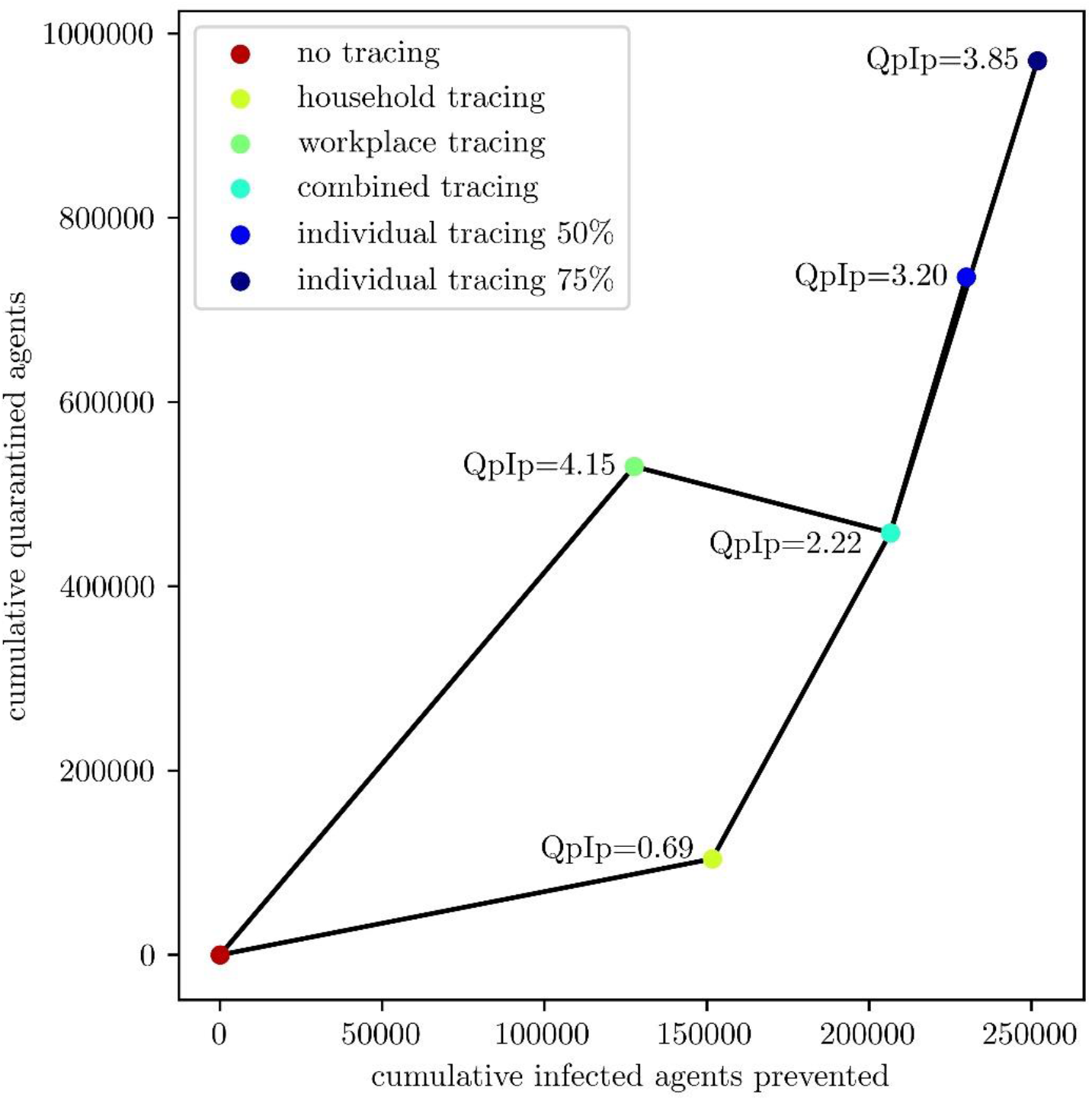
*Cumulative quarantined* agents plotted against *cumulative infected* agents prevented by tracing compared to the no tracing scenario. The valued displayed corresponds to the defined cost measure *quarantined per infection prevented (QpIp)*.

**Figure 7:**
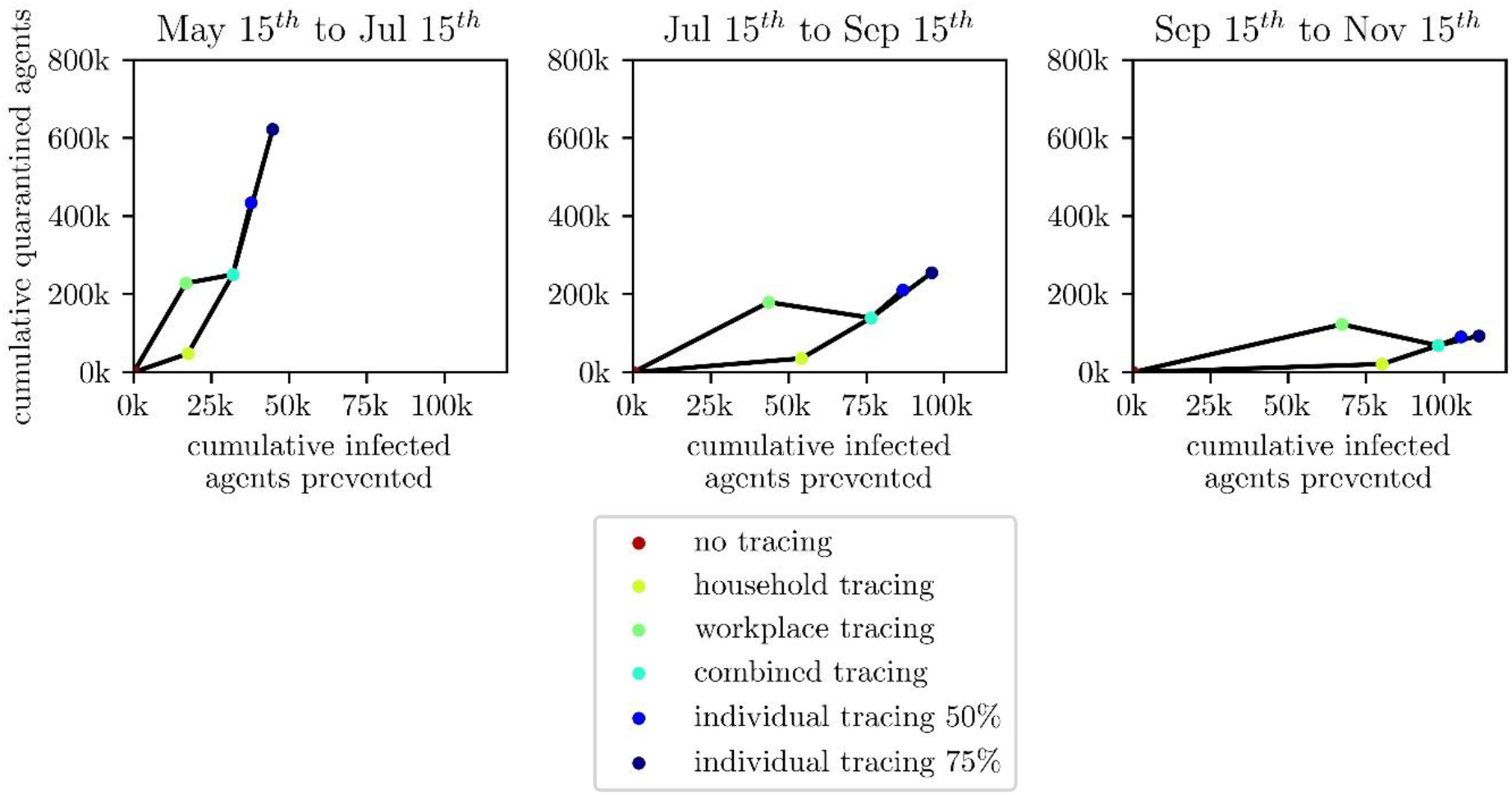
Analogous curves to Figure 6, yet the cumulative cases are not evaluated for the whole period May 15^th^ to Nov 15^th^, but for three sub-intervals with two months each.

## 4 Discussion

We implemented an agent-based simulation model that can not only be calibrated to match the previous course of the COVID-19 epidemic wave in Austria but is also capable of making comparisons between various non-pharmaceutical intervention strategies. In this study we applied this model to compare tracing policies in different characteristics and evaluated them with respect to socioeconomic costs in form of preventively quarantined people.

### 4.1 Evaluation of Tracing Policies

The model results indicate that tracing, in any form, is a suitable policy to contain the disease and can supplement lockdown policies if and only if combined with additional contact reduction. The results depicted in Table 1 indicate that well performed contact tracing could supplement for up to 5.18% contact or infectivity reduction. In contrast to required 75% reduction for full containment of the disease, this would only correspond to one small part of the full containment strategy. Yet, since tracing is one of the few strategies that does not impact the personal life of the population, such as closure of schools or limitation of movement, it must not be underrated. Note that all of the presented strategies are applied on top of a classical quarantine strategy in which any positively tested person is isolated. This is, essentially, the basis of any containment strategy and therefore a persistent element of the base model.

Anyway, isolating persons due to a preventive quarantine measure is always related to unintended economic and sociopsychological adverse effects, which is particularly critical if the isolation turns out to be unnecessary. Consequently, any tracing measure should focus on keeping the total number of isolated persons as small as possible to reduce socioeconomic damage. The defined cost value *Qp*I*p* is used to quantify the efforts of a specific tracing strategy. It relates with the direct benefit of the policy and directly correlates with the accuracy of the measure, that is the probability that a preventively isolated person is not only potentially but actually infected. Thus, the model suggests that isolation of household members is the most accurate measure and leads to the highest number of infections averted in relation to quarantined persons. Temporary closing of workplaces due to positive cases is clearly the least accurate and therefore the costliest of the modelled policies. Combining the two policies and adding additional leisure-time contact reduction also results in a more costly strategy, yet more infectivity reduction can be supplemented since more secondary infections are found and isolated. The model results show that tracing might require up to four times as many quarantined as infected persons for the least accurate tracing method (individual tracing with 75%) and about 0.7 times as many for the most accurate tracing method (preventive household quarantine). Considering that PCR tests used for detection of the index case are not 100% specific in reality, false-positive cases would be found and traced contacts would also be put falsely under quarantine. Since our model does not include this mechanism the ratio between quarantined and infected and also the *QpIp* values would be slightly higher in the real system than displayed above, dependent on the specificity of the test and the prevalence in the tested cohort.

In summary, the model results imply that a tracing strategy can be evaluated in terms of **effectiveness**, that is how much contact reduction can be supplemented by the strategy, and in terms of **accuracy**, that is how many persons need to be quarantined in relation to the averted infections, and that there is a tradeoff between these values – at least for the tracing strategies observed in this work.

Moreover, Figure 7 and Table 3 suggest that an inaccurate tracing method might also pay off in the long run if it is an effective one. This is due to highly interesting dynamics caused by the interplay of the two feedback loops depicted in Figure 8. If the feedback loop of the infectious persons dominates the system, a lot of new infections will increase the number of persons in preventive isolation and therefore the economic costs. Increasing the strictness of the tracing measure, that is trace more rigorously, will contribute to make the right feedback loop dominant and contain the disease. Yet, it directly increases the number of quarantined people at first. If applied in a successful containment strategy, both the infected and the preventively isolated people can be kept on a low level.

**Figure 8:**
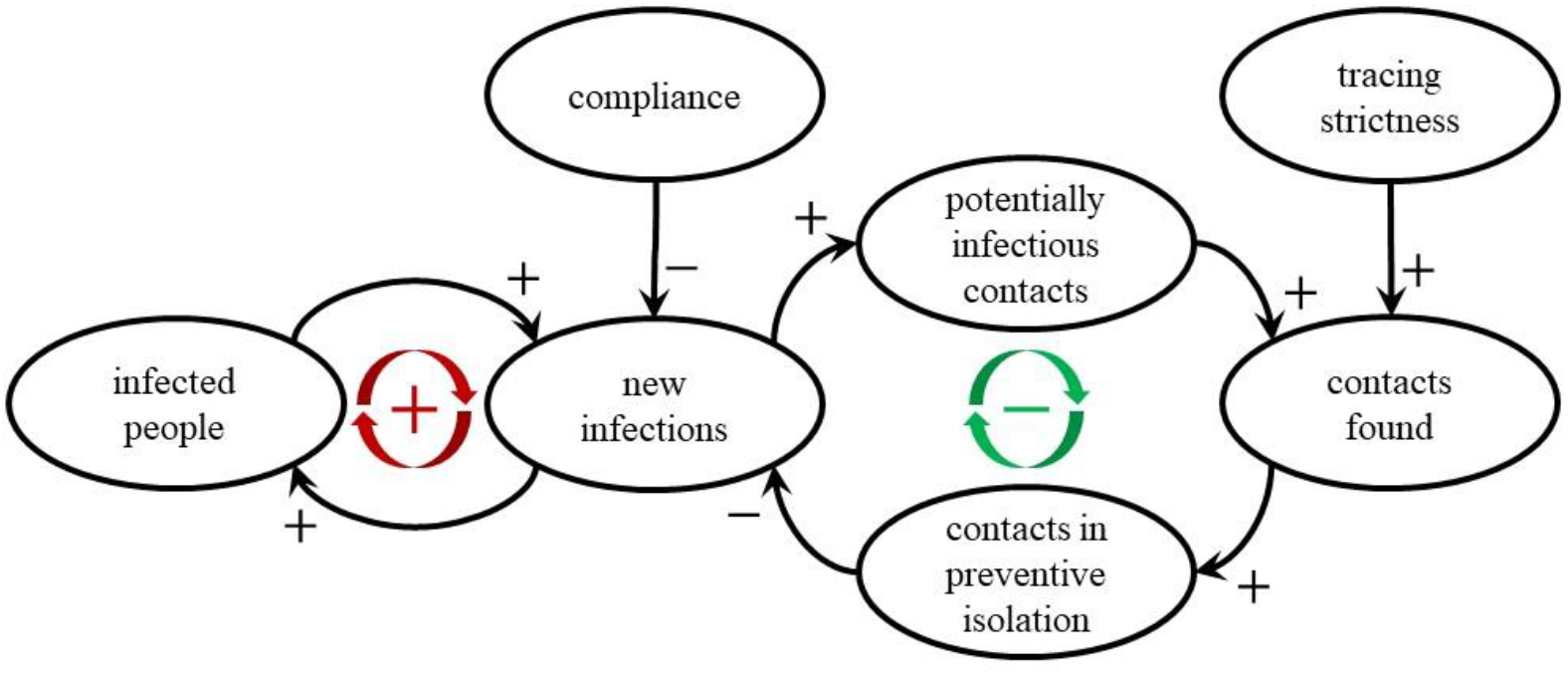
Causal Loop Diagram of relevant system components with respect to tracing measures. The more dominant the feed-back loop on the left-hand side, the more potentially infectious contact partners need to be isolated to contain the disease.

The model results nicely display the impact of this feedback in Figure 7 and Table 3. While peaking the increasing case numbers in May and June requires lots of preventive quarantined persons, the low case numbers in the well contained disease from July to November reduce the costs for the strategy. Consequently, the model results support the idea that tracing is much more efficient when case numbers are low – not only due to limited tracing resources. Hence, a strict contact tracing policy is costly in the first place but pays off in the long run.

In general, the findings of this work match the experience of countries that already implemented large-scale and very accurate contact tracing like Singapore or South Korea (31,32) and had great success with the strategy. However, both countries have already experienced that pure contact tracing alone is not sufficient to fully contain the disease.

Moreover, recent results from Austria contribute to the validation process of this study. While the country managed to contain the case numbers during the summer months, a second wave started to become eminent in October when the case numbers started growing with *R*_*eff*_∼1.2. On October 20^th^ the case numbers hit 1500 new confirmed cases per day and the ascend almost instantaneously steepened to a higher level of *R*_*eff*_∼1.4 (33).

This increase by 0.2 came surprising to Austrian decision makers since there was neither any policy, holiday nor weather change involved that might have explained it. Yet, having a closer look on cluster data (33) indicated that the Austrian contact tracing regime, although never officially confirmed, might have run out of resources at this time.

In summary, the model indicates that all tracing measures can supplement a small amount of infectivity reduction and can therefore play a role in a successful containment strategy. This strategy is particularly valuable since it does not require additional behavior restrictions from the common public, but only focuses a part of the contact network of positively tested persons. The related costs can be measured in preventively quarantined persons which are not only reduced by accurate tracing but also by low case numbers.

### 4.2 Limitations

We distinguish between limitations of the study design, the limitations of the simulation results and the limitation of the model as a decision support tool.

First of all, the study design does not include any testing of detected contact persons. Finding additional positive cases within contact partners would increase the capabilities of the strategy to enhance tracing to the next generation and would slightly increase the efficacy of the policy. Moreover, the study does not specifically regard sensitivity or specificity of the test that is used to determine the index cases for contact tracing. Test sensitivity is implicitly depicted in the general detection probability of COVID-19 cases. Test specificity is not regarded at all which was already analyzed in the discussion section. Moreover, the duration between start of infectivity and receiving of the test result is parametrized for typical PCR tests and might be different for antigen tests or others. The reader is referred to the parameter tables in the Appendix for more information.

The model results are, as all simulation outcomes, limited by modeling and data uncertainty: Many disease parameters of the novel coronavirus are still unknown and will surely improve in future due to increasing availability of data. In particular, the model suffers from the reporting bias of the calibration data. The constantly changing and limited availability of tests in the first months of the disease might cause the data to estimate the real epidemiology falsely.

Moreover, the parameters and the modeling assumptions for the tracing strategies are based on expert opinions. Consequently, the tracing algorithm is modeled in an oversimplified form and cannot validly depict the actual work of professional contract tracers.

Interpreted as a decision support tool, the model is primarily limited by comparably long computation times and fundamental simplifications made during the modeling process. The prior is caused by the problem that the model cannot be scaled down and always requires simulation with the full population of the country. Thus, a huge number of agents (about 9 Million for Austria) leads to long computation times, and the necessity of Monte Carlo simulation for flattening of stochastic results increases the time required to get simulation output even more. Consequently, the simulation’s capabilities of dealing with multi-variate calibration problems are limited and the model is capable, yet unhandy to generate short-time prognoses.

### 4.3 Conclusion

We presented an agent-based simulation model that is not only capable of simulation of epidemics but can also be used to evaluate and compare different containment strategies: Beside classic lockdown interventions like closure of schools or workplaces, the model can also be used to compare different tracing policies which makes it unique and powerful.

Hereby, we also displayed the limits of classical cohort models, as comparable scenarios would not be feasible with aggregated modeling approaches: By aggregating individual contacts into global contact rates, individual contact-chains are lost, and tracing cannot be modelled.

For our policy question on tracing, we investigated six tracing strategies for three compliance scenarios regarding a second outbreak of COVID-19 in Austria. They allowed us to simulate and quantify the impact of different tracing policies and draw conclusions about tracing in general.

The results demonstrate that tracing of potentially infectious contacts and subsequent isolation of affected persons is a very useful measure to slow the spread of SARS-CoV-2 and that there are more and less restrictive ways to do so. The model results display that a well contained disease also reduces the socioeconomic costs for tracing in terms of fewer quarantined persons. Consequently, the model results recommend strict and accurate tracing strategies in favor of wide range preventive closure of locations like workplaces which cause many unnecessarily quarantined healthy persons. Yet, the model results also show that tracing in any variant, though effective, can only play a minor role in disease containment.

Evaluating the effectiveness of tracing policies is only one of many features of this advanced ABM. Although the model has limitations, it is a well-founded basis for COVID-19 related decision support as it can include complicated model-logic and diverse and high-resolution data. Hence, we plan to extend our policy comparisons started in this study in the future by direct comparison of other interventions, such as a more advanced testing regime or the introduction of a vaccine.

## Supporting information

Appendix (Model Description)

## Data Availability

All parametrisation data and links to data sources are found in Appendix and References.

## Footnotes

1 This number corresponds to the actual state of the confirmed cases on the specified date at the specified time. Due to a reporting bias, this number is subject to constant changes and will probably increase in the future.

## References

1. Organization WH, others. Coronavirus disease 2019 (COVID-19): situation report, 88. 2020;

2. Ferrazzi EM, Frigerio L, Cetin I, Vergani P, Spinillo A, Prefumo F, et al. COVID-19 Obstetrics Task Force, Lombardy, Italy: Executive management summary and short report of outcome. Int J Gynecol Obstet. 2020 Jun;149(3):377–8.

3. Grasselli G, Pesenti A, Cecconi M. Critical Care Utilization for the COVID-19 Outbreak in Lombardy, Italy: Early Experience and Forecast During an Emergency Response. JAMA. 2020 Apr 28;323(16):1545.

4. Daily C. Three steps for Wuhan to achieve “beds waiting for patients.” wuhan. gov. cn. 2020.

5. Nicola M, Alsafi Z, Sohrabi C, Kerwan A, Al-Jabir A, Iosifidis C, et al. The socio-economic implications of the coronavirus pandemic (COVID-19): A review. Int J Surg. 2020 Jun;78:185–93.

6. Cho H, Ippolito D, Yu YW. Contact tracing mobile apps for COVID-19: Privacy considerations and related trade-offs. ArXiv Prepr 200311511. 2020;

7. COVID-Prognose-Konsortium [Internet]. Available from: https://www.sozialministerium.at/Informationen-zum-Coronavirus/COVID-Prognose-Konsortium.html

8. Kermack WO, McKendrick AG. A Contribution to the Mathematical Theory of Epidemics. Proc R Soc Math Phys Eng Sci. 1927 Aug;115(772):700–21.

9. Macal CM, North MJ. Tutorial on Agent-Based Modeling and Simulation Part 2: How to Model with Agents. In: Proceedings of the 2006 Winter Simulation Conference [Internet]. Monterey, California; 2006 [cited 2010 Apr 12]. p. 73–83. Available from: http://www.informs-sim.org/wsc06papers/008.pdf

10. Macal CM, North MJ. Agent-based Modeling and Simulation. In: Proceedings of the 2009 Winter Simulation Conference [Internet]. New York, N.Y.: Association for Computing Machinery; 2009 [cited 2011 Aug 30]. p. 86–98. Available from: http://www.informs-sim.org/wsc09papers/009.pdf

11. Caro JJ, Briggs AH, Siebert U, Kuntz KM. Modeling Good Research Practices—Overview: A Report of the ISPOR-SMDM Modeling Good Research Practices Task Force-1. Value Health. 2012 Sep;15(6):796–803.

12. Pitman R, Fisman D, Zaric GS, Postma M, Kretzschmar M, Edmunds J, et al. Dynamic Transmission Modeling: A Report of the ISPOR-SMDM Modeling Good Research Practices Task Force Working Group–5. Med Decis Making. 2012 Sep;32(5):712–21.

13. Chang SL, Harding N, Zachreson C, Cliff OM, Prokopenko M. Modelling transmission and control of the COVID-19 pandemic in Australia. ArXiv. 2020;abs/2003.10218.

14. Ferguson N, Laydon D, Nedjati Gilani G, Imai N, Ainslie K, Baguelin M, et al. Report 9: Impact of non-pharmaceutical interventions (NPIs) to reduce COVID19 mortality and healthcare demand. 2020;

15. Mossong J, Hens N, Jit M, Beutels P, Auranen K, Mikolajczyk R, et al. POLYMOD social contact data. 2017;

16. Web-page of the City of Vienna [Internet]. Available from: https://www.wien.gv.at/

17. Austria WS. Arbeitsstättenzählung 2001. Verlag Österreich; 2004.

18. Austria S. Bildung in Zahlen 2017/18-Tabellenband. 2019;

19. Hellewell J, Abbott S, Gimma A, Bosse NI, Jarvis CI, Russell TW, et al. Feasibility of controlling COVID-19 outbreaks by isolation of cases and contacts. Lancet Glob Health. 2020;

20. Lauer SA, Grantz KH, Bi Q, Jones FK, Zheng Q, Meredith HR, et al. The incubation period of coronavirus disease 2019 (COVID-19) from publicly reported confirmed cases: estimation and application. Ann Intern Med. 2020;

21. COVID-19 information page by AGES [Internet]. Available from: https://www.ages.at/en/wissen-aktuell/publikationen/epidemiologische-parameter-des-covid19-ausbruchs-oesterreich-2020/

22. Gudbjartsson DF, Helgason A, Jonsson H, Magnusson OT, Melsted P, Norddahl GL, et al. Spread of SARS-CoV-2 in the Icelandic Population. N Engl J Med. 2020 Jun 11;382(24):2302–15.

23. Zauner G, Popper N, Breitenecker F. State of the Art Research In Austria: Dexhelpp - Decision Support For Health Policy and Planning: Methods, Models and Technologies Based On Existing Health Care Data. Value Health. 2014 Nov;17(7):A452.

24. Bicher M, Urach C, Popper N. GEPOC ABM: A Generic Agent-Based Population Model for Austria. In: Proceedings of the 2018 Winter Simulation Conference. Gothenburg, Sweden: IEEE; 2018. p. 2656–67.

25. Grigoryev I. AnyLogic 6 in three days: a quick course in simulation modeling. [Hampton, NJ]: AnyLogic North America; 2012.

26. Tisue S, Wilensky U. NetLogo: A simple environment for modelling complexity. In 2004. p. 16–21.

27. Masad D, Kazil J. MESA: an agent-based modeling framework. In: 14th PYTHON in Science Conference. 2015. p. 53–60.

28. Bellifemine F, Poggi A, Rimassa G. JADE–A FIPA-compliant agent framework. In: Proceedings of PAAM. London; 1999. p. 33.

29. North MJ, Howe TR, Collier NT, Vos JR. The repast simphony runtime system. In: Proceedings of the agent 2005 conference on generative social processes, models, and mechanisms. Citeseer; 2005. p. 13–5.

30. dwh GmbH news entry for the ABT simulation framework [Internet]. Available from: http://www.dwh.at/en/news/the-power-of-the-abt-simulation-framework/

31. Moradi H, Vaezi A. Lessons learned from Korea: COVID-19 pandemic. Infect Control Hosp Epidemiol. 2020;1–2.

32. Pung R, Chiew CJ, Young BE, Chin S, Chen MI, Clapham HE, et al. Investigation of three clusters of COVID-19 in Singapore: implications for surveillance and response measures. The Lancet. 2020;

33. Richter L, Schmid D, Chakeri A, Maritschnik S, Pfeiffer S, Stadlober E. Epidemiologische Parameter des COVID19 Ausbruchs–Update 08.01. 2021, Österreich, 2021. Update. 2021 Jan 8;3(2021).

