## Appendix (Model Description) for "Evaluation of Contact-Tracing Policies Against the Spread of SARS-CoV-2 in Austria– An Agent-Based Simulation"

### A1 Detailed Model Specification

We will explain our agent-based COVID-19 model based on the ODD (Overview, Design Concepts, Details) protocol by Volker Grimm et.al. [22, 23].

#### A1.1 Overview

The modelling of the spread of the disease is based on the interplay of four modules.

1. Population. Altogether the agent-based COVID-19 model is based on the Generic Population Concept (GEPOC, see [14]), a generic stochastic agent-based population model of Austria, that validly depicts the current demographic as well as regional structure of the population on a microscopic level. The flexibility of this population model makes it possible to modify and extend it by almost arbitrary modules for simulation of population-focused research problems.

2. Contacts. In order to develop a basis for infectious contacts, we modified and adapted a contact model previously used for simulation of influenza spread. This model uses a distinction of contacts in different locations (households, schools, workplaces, leisure time) and is based on the POLYMOD study [29], a large survey for tracking social contact behaviour relevant to the spread of infectious diseases.

3. Disease. We implemented a module for the course of the disease that depicts the current pathway of COVID-19 patients starting from infection to recovery or death and linked it with the prior two modules.

4. Policies. Finally, we added a module for implementation of interventions, ranging from contact-reduction policies, hygienic measures and, in particular, contact tracing. This module is implemented in form of a timeline of events.

##### A1.1.1 Purpose

The agent-based COVID-19 model aims to give ideas about the potential impact of certain policies and their combination on the spread of the disease, thus helping decision makers to correctly choose between possible policies by comparing the model outcomes with other important factors such as socioeconomic ones. In order to fulfil this target, it is relevant that the agent-based COVID-19 model validly depicts the current and near future distribution and state of disease progression of infected people and their forecasts.

In the following overview of the model, we will not state any parameter values to focus on the model concept. A full collection of model parameters including values, sources and justifications is found in Section A1.3.3.

##### A1.1.2 Entities and State Variables

Each person-agent is a model for one inhabitant of the observed country/region. We describe state variables of a person-agent sorted by the corresponding module.

Population. Each person-agent contains the population specific state variables *sex*, *date of birth* (≅ age) and *location.* The latter defines the person-agent's residence in form of latitude and longitude and uniquely maps to the agent's municipality, district and federal state.

Contacts. The person-agent features a couple of contact network specific properties. These include a *household* and might include a *workplace* or a *school.* We summarise these as so-called *locations* which stand for network nodes via which the person-agent has contacts with other agents. Assignment of person-agents to *locations* is based on distance of the agent's residence to the position of the *location.* Each day, an agent has a certain number of contacts within each of the *locations*, which essentially leads to spread of the disease. To model contacts apart from these places, every person-agent has an additional amount of leisure time contacts, which are sampled randomly based on a spatially-dependent distribution. The contact network is schematically displayed in Figure A1.

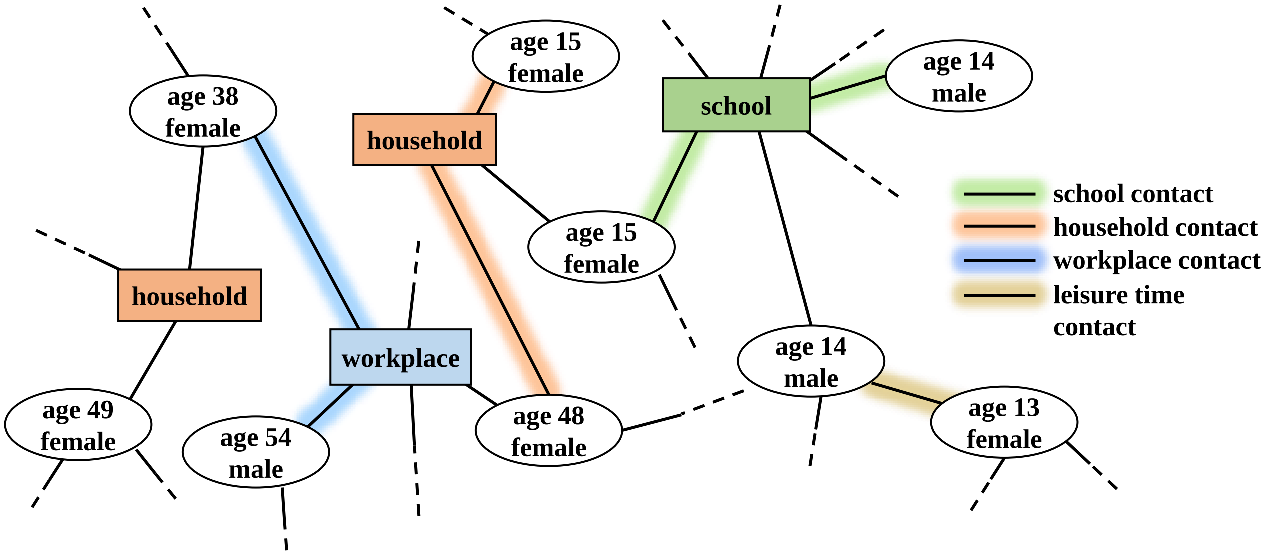


Figure A1: Contact network of agents in the agent-based COVID-19 model. Regular contacts between agents occur via locations (schools, workplaces and households, while random leisure time contacts extend the standard contact network.

Disease. In order to model the spread of the disease each person-agent has a couple of health states that display the current status of the agent. The parameter necessary for the simulation dynamics are *susceptible*, *infected*, *infectious*, *confirmed*, *hospitalised*,*quarantined* and *critical*. These states can either be true or false, and multiple of them can be true at a time. They stand for certain points within the patient pathway of an infected person and enable or disable, respectively, certain person-agent actions. The meaning of these attributes is usually self-explanatory: for example an agent with attribute *confirmed=true* indicates that the agent has been detected by by a SARS-CoV-2 test. To make generation of simulation output easier, we sometimes make use of derived parameters such as *severe* ($=hospitalized \wedge \neg critical$) or *undetected* ($=infected \wedge \neg confirmed$). More on the influence of these state variables and how they change is described in Section A1.1.4.

Policies. Policies apply either to *locations* or to person-agent-behaviour directly and require additional agent properties. All *locations* except for households are defined *open* or *closed* which marks whether this place is available for having contacts. For person-agents the variable *quarantined* is applied to mark agents isolated not only via a positive test but also due to tracing measures.

For the sake of simplicity of speech we furthermore address mentioned parameters as attributes for the corresponding agents. I.e. an agent with *infectious* set to true will be denoted as “*infectious* agent”.

##### A1.1.3 Scales

Unlike other agent-based models it is not possible to validly run the model with a smaller number of agents (e.g. one agent represents 10 or 100 persons in reality) as certain contact-network parameters do not scale this way (average school size,...). Consequently, one simulation run always uses agents according to the size and structure of the full population.

##### A1.1.4 Process Overview and Scheduling

Like the underlying population model, the agent-based COVID-19 model can be interpreted as a hybrid between a time-discrete and a time-continuous (i.e. event-updated) agent-based model:

The overall simulation updates itself in daily time steps, wherein each step is split into three phases. In the first phase each agent is called once to plan what it aims to do in the course of this time step. In the second phase, each agent is, again, called once to execute all planned actions for this time step in the defined order. In the third step, a recorder-agent keeps track of all aggregated state variables.

On the microscopic scope, each person-agent is equipped with its own small discrete event simulator. In the mentioned planning phase, each agent schedules certain events for the future which may, but not necessarily must, be scheduled within the current global time step. In the second phase, the agent executes all events that are scheduled for the currently observed time interval, but leaves all events that exceed this scope untouched.

This strategy comes with the following benefits:

- In contrast to solely event-based ABMs, the event queue is distributed among all agents which massively increases the speed for sorting (a solely event-based ABM with millions of complex agents would not be executable is feasible time).
- Moreover, in contrast to solely event-based ABMs, usage of daily transition probabilities/rates instead of transition times is possible as well.
- In contrast to solely time-discrete ABMs, agents can operate beyond the scope of time steps and sample continuous time-intervals for their state-transitions.

We shortly describe all actions that are scheduled and executed by one person-agent within one time step sorted by the specified module.

Population. As briefly described in [14], agents trigger birth and death events always via time- and age-dependent probabilities that apply for the observed time step (i.e. the observed day). If one of these events triggers, the agent samples a random time instant within the current time step and schedules the event. Note that in contrast to the basic population model, immigration and emigration events are disabled in the agent-based COVID-19 model due to closed borders in reality.

Contacts. Also contact specific events are scheduled and executed within the scope of only one time step: First of all, the agent schedules a contact event with every other member of its *household.* Moreover, if such a location is present in the contact network, a certain number of *workplace* or *school* contacts, respectively, are scheduled and corresponding partners drawn randomly from the assigned location. Finally, a certain number of *leisure time* contacts are sampled and partners are drawn based on an origin-destination matrix on municipality resolution. The latter has been gathered from mobile data (see Tables A2-A3).

As mentioned, some states limit the agents' capabilities of interaction. In specific, *quarantined* agents have no random leisure time contacts and no contacts at *work* or *school.* Furthermore, *hospitalised* agents do not even contact their household members.

Anyway, planned contacts are always scheduled for the beginning of the new time-step. Hence, interaction between agents is actually limited to the discrete time steps of the simulation. This guarantees, that the states of both involved agents do not differ between the time of the planning of the event and its execution.

Disease. First of all, it is important to mention that the model is not parametrised by a reproduction number $R_0$ or $R_{eff}$, but by a contact-specific probability for a transmission in case of a contact. Nevertheless, the agent-based model provides the opportunity to generate estimates for $R_0$ and $R_{eff}$ by its original definition: the average number of secondary infections of an infected agent. Hence, what comes as model input for many traditional SIR models becomes a model output for the agent-based COVID-19 model.

In case of a contact, *infectious* agents spread the disease to *susceptible* agents with a certain *infection probability* which triggers the start of the newly-infected agent's patient-pathway. This pathway describes the different states and stations an agent passes while suffering from the COVID-19 disease and can be interpreted as a sequence of events of which each triggers the next one after a certain sampled duration.

We show this infection strategy in a state chart in Figure A2 and describe how to interpret this figure by explaining the initial steps in the pathway in more detail: As soon as a person-agent becomes infected, its *infected* state is set to *true*, its *susceptible* variable is set to *false* and a *latency period* is sampled according to a specific distribution. The corresponding “Infectious” event is scheduled for the sampled time instant in the future. As soon as this “Infectious” event is executed, the *infectious* parameter is set to *true* and a random number decides about whether the person will develop symptoms or not. This point marks the first branch in the patient’s pathway and whether the “Symptoms Onset” event or the “Unconfirmed Disease” Event is scheduled. The prior would be planned after a sampled time span corresponding to the difference between latency and incubation time, the latter would be triggered instantaneously. All other elements of the pathway follow analogously. The branches are evaluated with age-class-dependent probabilities (see Section A1.3.3).

In most cases (i.e. if the agent does not die for any other non-COVID related reason - see Population module), the final state of every agent’s disease pathway is the *Recovery/Removal* event with either sets the agent *resistant*, or renders it deceased with a certain death-by-COVID probability that depends on the agent’s disease severity. Consequently, the model differs between COVID-caused and COVID-affected deaths.

Policies. Every policy is modelled as a global event occurring before the planning phase of any of the simulation time steps. Policies are timed-events that are fed into the model as an event-timeline (see Figure A3). The elements of this timeline may include real policies like closure or opening of locations, start of tracing (for a full list, see Table A7), but may also contain incidents that change the model behaviour but are not directly related to policies, such as raising hygiene awareness or distancing. The most outstanding feature of the model is clearly its ability to model contact tracing policies, since agents are aware of all other agents with which they had contacts. Using simple housekeeping arrays, these can be logged for a certain period of time and used for detection and isolation of contact partners.

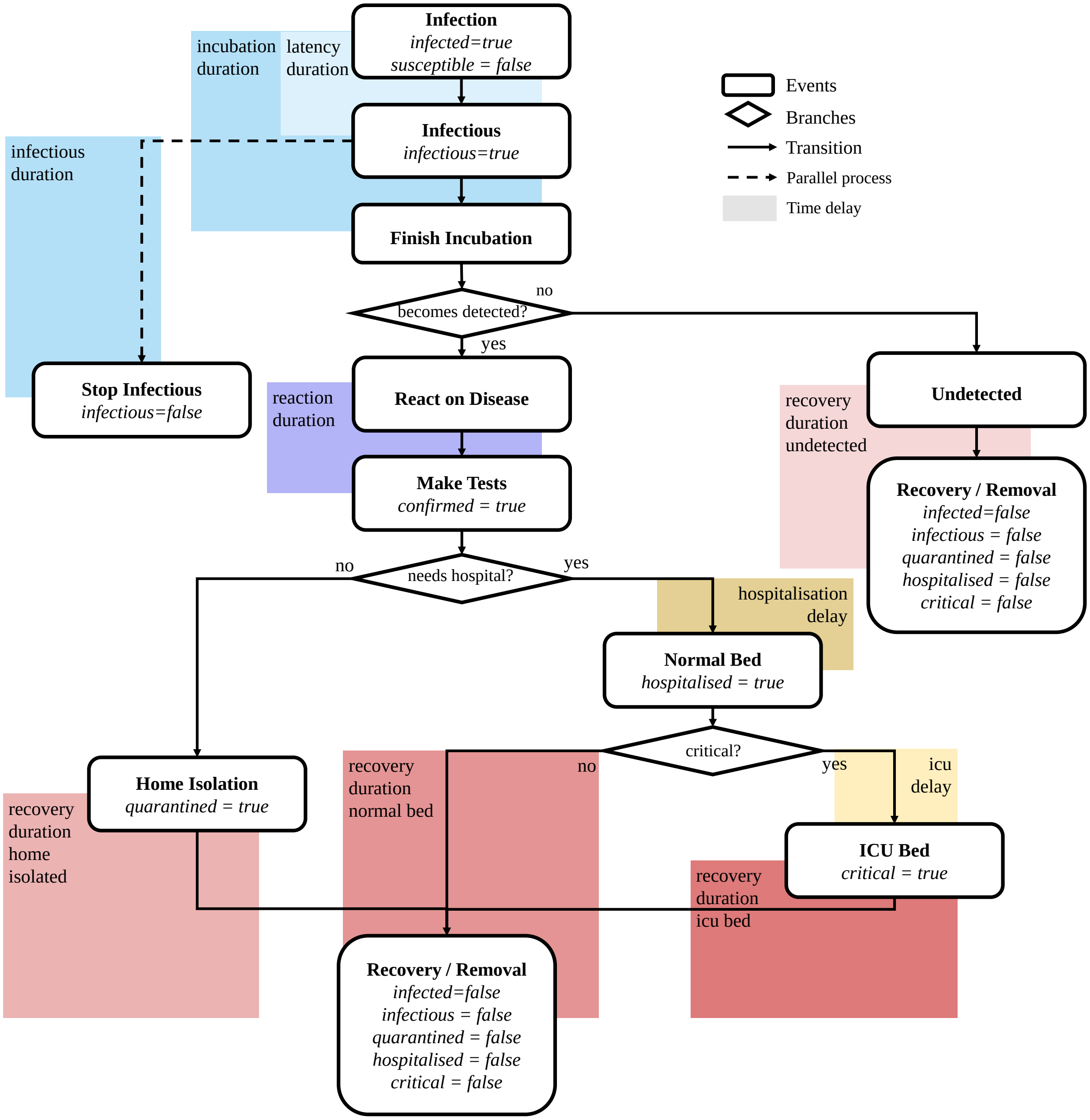


Figure A2: State chart of the patient pathway of a person-agent in the agent-based COVID-19 model. Only those state variables that are changed by the corresponding event are labelled, all others remain at the current value. The initial state of all infection-specific state variables is *false*, except from *susceptible* which is initially *true*.

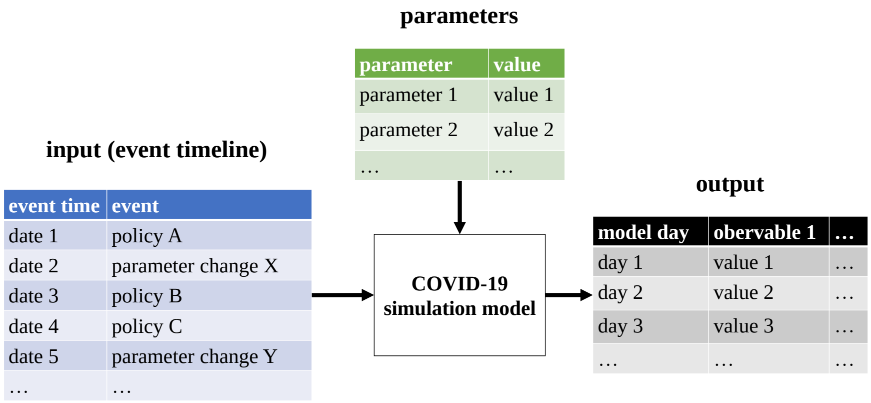


Figure A3: Event-timeline as the input of the simulation model in contrast to standard model parameters.

#### A1.2 Design Concept

##### A1.2.1 Basic Principles.

Increasing the level of detail from a standard epidemiological model for simulation of disease waves to a model that is capable of dealing with various different policies is a huge step with respect to model complexity. It excludes the use of macroscopic strategies and requires modelling of a contact network and contact behaviour. Consequently a detailed demography, spatial components and stochasticity need to be introduced to the model which come with a huge number of additional parameters and parameter values.

Hence, we were very careful that the agent-based model is designed as simple as possible yet tracking the most important features for evaluation of certain policies. Hereby, many details within the pathway of an infected person and, in particular, lots of details within the personal daily routine are simplified to avoid indeterminable model parameters and unpredictable model dynamics.

##### A1.2.2 Emergence.

In addition to the classic emergence of nonlinear epidemiological effects, analysis of the effects of interaction between different measures is one of the key objectives of the model. For example, seemingly unconnected policies like *school closure* and *contact reduction for the 65+* might lead to unexpected effects when applied simultaneously. More generally speaking, the model displays that the individual effects of applied policies do not add up linearly.

##### A1.2.3 Sensing.

Agents' perception of reality is one of the key problems of modelling COVID-19 as no agent is actually aware of its own disease and, more importantly, infectiousness until symptoms occur. Therefore, agent parameters can be distinguished into two sets: the ones the agent is aware of (e.g. *critical*, *hospitalised*), and the ones it is not (e.g. *infected*, *infectious*).

Interestingly, besides the individual perception of agents and the perception of an omniscient observer, there is also a third level of perception included into the model: the perception of the general public. While an individual agent knows about its symptoms, the public is not yet aware of this additional infected case, until the person-agent has reacted on the disease, has had itself tested and eventually becomes *confirmed*.

Consequently, the levels of perception can be sorted with regards to their amount of knowledge:

$$\text{omniscient observer} > \text{person-agent} > \text{general public}.$$

###### A1.2.4 Interaction.

Interaction between agents only occurs in form of contacts at *locations* or *leisure time.* The features provided by the underlying population model make it possible to investigate contacts on a very local level. As described before, *leisure time* contacts are weighted by their regionality, but also *school* and *workplace* contacts depict locality: Using specified latitude and longitude for locations, it is possible to assign person-agents with distance-dependent probabilities (see Section A1.3.1). Consequently, interactions between agents follow a spatially-continuous locally-biased contact network.

###### A1.2.5 Stochasticity.

Basically all model processes, including the initialisation, contain sampling of random numbers. Therefore, Monte Carlo simulation is applied, results of runs are averaged and also their variability is assessed (see Section A3).

###### A1.2.6 Observation.

Inspired by [30], a recorder-agent takes care about tracking and aggregating the current status of the simulation. At the end of each global time step, all person-agents report to the recorder-agent which furthermore keeps track of all necessary aggregated model outputs. This includes for example *confirmed active cases*, *confirmed cumulative cases*, *hospitalised agents*, *undetected agents*, *pre-symptomatic agents*, *recovered agents*, *agents in a certain hospital*, or *average-number of contacts per infectious agent.* If required, numbers can also be tracked with respect to age, sex, regional level and/or location.

##### A1.3 Details

Clearly, Section A1.1 could only outline the basic concepts of the model and left a lot of technical and modelling details necessary for a reproducible model definition open. In particular, this refers to the highly non-trivial initialisation process of the model. Hereby, two problems occur that require completely different approaches. The first problem considers the generation of the person-agents, locations and hospitals in the first place. The second problem deals with the initialisation of the status quo of the distribution of the disease states of the agents for the specified initial date.

###### A1.3.1 Initialisation of Person-agents, Locations and Hospitals.

A lot of problems that deal with the sampling of the initial population have already been solved in the original GEPOC model [14]. In particular this refers to the Delaunay-triangulation-based sampling method for locations. We apply this method to merge information from the national statistics institute and the global human settlement layer [20]. Consequently, besides initialisation of the disease states which is described in the next section, only new methods for location- and hospital-generation had to be implemented.

Schools are initialised based on known distributions w.r.t. average school size and number of pupils in total. A *school-sampler* iteratively generates schools with a random size/capacity (truncated normal distribution) until the sum of all capacities matches the known number of pupils in reality. Each school is furthermore sampled a position (latitude and longitude) analogous to the sampling for person-locations (see [14]). In a second step, schools are "filled" with person agents. Hereby, model agents with age between 6 and 18 are assigned to a school based on a locally biased distribution. This is done with probabilities $p^s_m, p^s_d, p^s_f$ and $p^s_a$ for assigning the school in the same municipality, district, federal-state and country as the agent's residence.

Clearly, the number of model agents in this age group is larger than the number of known pupils. Consequently, we force distribution of all 6 to 14 year old agents, and distribute as many 15 to 18 year old agents as possible. All remaining 15 to 18 year old agents are considered to be working.

Workplaces[1] are initialised analogously to schools. A *workplace-sampler* iteratively generates workplaces with size/capacity according to a discrete distribution (see Tables A2-A3). The sampler stops generating if the sum of all capacities matches $(a + b)(1 - \alpha) - c$, whereas $a$ denotes all model agents between 19 and 64, $b$ denotes all agents between 15 and 18 that have not been yet assigned a school, $c$ denotes the total number of care-home workers in Austria, and $\alpha$ denotes the current unemployment rate. Location sampling and "filling" works analogously to the *school-sampler* with different probabilities (commuter data).

Care-homes are generated with a fixed size and providing space for a fixed number of inhabitants and workers ($= c$, see above). Workers and inhabitants are assigned based on the same distribution as workplaces, although assigning inhabitants actually transfers theirs residence to the sampled place of the *care-home*.

Hospitals are generated based on publicly available data. This includes capacities (beds, intensive-care units) as well as their location (latitude and longitude).

###### A1.3.2 Initialisation of the Disease State

The spread of SARS-CoV-2 displays probably better than any other system, that the most dangerous enemy is the invisible one. While confirmed infected persons are detected and well known, they hardly contribute to the spread of the disease – they are already isolated properly, and most infections occur even before the onset of symptoms.

Consequently, it is not possible to simply "start" the simulation with a certain number of confirmed cases, acquired for example from official internet sources. Valid values for pre-symptomatic (e.g. persons within latency and incubation period) and asymptomatic persons need to be acquired as well – yet, this number is hardly measurable in reality.

In order to solve this problem, a three stage concept, henceforth denoted as initialisation phase, was designed to generate a feasible initial state for a certain time $t_0$:

1. **Initialise-Simulation.** The agent-based COVID-19 model is set up with a small number of initially infected agents (40 was found to be the most stable and useful option). This number corresponds to an estimated count of initial infection clusters in the country, but actually hardly influences the outcome. Furthermore, the agent-based simulation is run and interrupted by a state event, namely if the cumulative number of *confirmed agents* in the model is greater or equal to a specific value $C(t_{-1})$. Hereby, $t_{-1}$ refers to a self chosen point in time and $C(t_{-1})$ to the reported number of positive tests in reality until $t_{-1}$. Hereby $t_{-1}$ must be chosen properly so that the reported number of positive tests is large enough to be representative yet before implementation of any policies.

   As soon as the simulation is interrupted by the state-event, the timelines of simulation and reality are synced: $t_{-1}$ in reality becomes $t_{-1}$ in the simulation.

   The initialise-simulation is continued, considering all policies that have been implemented in reality, until, finally, $t_0$ is reached. Properly calibrated by a calibration routine (see Section A1.3.4), the initialise-simulation contains approximately the same cumulative number of *confirmed agents* as the corresponding reported number in the real system.

   The initialise-simulation is finished by exporting parts of the final state of the simulation. This refers to all households that contain either infected or recovered agents which are finally written into a file. Hereby, an initial population is generated that contains not only a valid approximation of the confirmed cases, but also a valid estimate for the unknown pre-symptomatic and asymptomatic persons, a correct distribution of their future planned events and a correct household distribution as well.

[1]Workplaces should not be confused with total companies. They rather represent the different teams where the members are in regular contact with each other.

2. **Fine Tuning.** Even with best calibration routines (see Section A1.3.4) it is not possible to perfectly match the model output with the status quo in reality, in particular w.r.t. regional distribution. Therefore, a bootstrapping algorithm was implemented that corrects the small differences between the initialise-simulation output and the real data (confirmed cases, hospitalisation, intensive-care units and recoveries per region) to make sure, that the initial state of the actual simulation matches the current state precisely. This step can be omitted, if matching the current state precisely is not required.

3. **Load Households.** Finally, the actual simulation is initialised with the previously recorded and fine-tuned agents from the initialise-simulation. To be precise, this process does not only include agents themselves, but also the *households* these agents live in. Hereby, the fundamental network structure from the initialise-simulation can be maintained.

##### A1.3.3 Parametrisation

With respect to parametrisation, we will distinguish between model input and model parameters.

Classical model parameters specify scalar or array-typed model variables that are initialised at the beginning of the simulation and, if not changed by certain model events, keep their value for the entire simulation time. Examples are the infection probability of the disease, the age-dependent death rate of the population, or the distribution parameters of the recovery time.

In contrast to model parameters, the model input consists of an event-timeline that describes at which point in time a certain incident changes the behaviour of the model. This incident usually refers to the introduction of a policy, e.g. closure of schools or start of tracing, but may also refer to instantaneous changes of model parameters which are related but cannot be directly attributed to policies, e.g. the increase of compliance among the population to increase hygiene.

In the following, we state lists of used parameters and parameter-values including corresponding sources and/or justifications. They are found in Tables A1 to A6. Table A7 states a list of possible event-timeline elements that can pose as the model's input.

Table A1: List of population specific parameters

| parameter | description | value | source |
|---|---|---|---|
| *birthrates*, *deathrates*, *initial population*, *regional distribution* | parameters used by the underlying population model | see source | rates and population tables from Austrian National Statistics Institute [8]. Maps from the Global Human Settlement Project [19] and [4]. |

##### A1.3.4 Calibration

Clearly, there is no valid data available for direct parametrisation of the *infection probability* in case of a direct contact. First of all, this parameter is hardly measurable in reality and moreover strongly depends on the definition of "contact". Consequently, this parameter needs to be fitted in the course of a calibration loop.

The calibration experiment is set up as follows:

- We vary the parameter *infection probability* using a bisection algorithm.
- For each parameter value, the simulation, parametrised without any policies, is executed ten times (Monte Carlo simulation) and the results are averaged.
- The average time-series for the *cumulative confirmed cases* is observed and cropped to the beginning upswing of the epidemic curve, to be specific, all values between 200 and 3200. In this interval the growth of the curve can be considered as exponential.

- The cropped time-series is compared with the corresponding time-series of real measured data in Austria, specifically the confirmed numbers between March 10$^{th}$ and 20$^{th}$ 2020 (source EMS system, [2]).
- Both time-series are compared w.r.t. the average doubling time of the confirmed cases. The difference between the doubling times is taken as the calibration error for the bisection algorithm.

Note: As the sample standard deviation of each observable of the runs has been observed to be at most a fifth of the sample mean, the iteration number of nine for the Monte Carlo simulation has been considered to be sufficient for calibration purposes w.r.t. the ideas in [15, 25].

After calibrating the infection probability parameter, also the parameters of the input timeline of the initialisation-simulation needs to be calibrated to the policies applied in the real system. This is done iteratively using a bisection algorithm as well. For example, the Austrian government introduced lockdown policies on March 17$^{th}$ and started with the first reopening steps on April 16$^{th}$. Consequently, this time-period was used to calibrate the unknown *leisure-time contact reduction* parameters in the lockdown-related policy events.

### A2 Model Implementation

Simulation of agent-based models like the agent-based COVID-19 model is a huge challenge with respect to computational performance. As the model cannot be scaled down, almost 9 Million interacting agents need to be included into the model in order to simulate the spread of the disease in Austria.

These high demands exclude most of the available libraries and software for agent-based modelling including AnyLogic [21], NetLogo [33], MESA [27], JADE [12] or Repast Simphony [31]. Most of these simulators cannot be used as their generic features for creating live visual output generates too much overheads.

Consequently, we decided to use our own agent-based simulation environment ABT (Agent-Based template, see [3]), developed in 2019 by dwh GmbH in cooperation with TU Wien. The environment is implemented in JAVA and specifically designed for supporting reproducible simulation of large-scale agent-based systems.

The next section contains more technical details about the implementation.

### A3 Technical Implementation Details

The implementation of the agent-based COVID-19 model uses JAVA 11 and applies the *UniformRandomProvider* random number generator (RNG) by Apache Commons [1]. This RNG implements a 64 bit version of the Mersenne Twister [28] and exceeds the standard RNG of JAVA, a simple Linear Congruential Generator, in both performance and quality.

The simulation itself is always executed in a Monte Carlo setting and several runs with different RNG seeds are averaged. Due to the huge number of agents, a Law-of-Large-Numbers-effect can be observed (similar to [13] Chapter 5.2), and the standard deviation of the model output is always comparably small. Consequently, Monte Carlo replication numbers of 10 to 20 are usually enough to estimate the mean sufficiently well (we apply the algorithms from [15, 25]).

### A4 Detailed Scenario Definition

In order to give a reproducible definition of scenarios, we explain the used event-timelines in detail in tables A8 and A9. The prior shows the calibrated timeline of initialization phase, the latter displays the timeline of the baseline scenarios.

This event-timeline was calibrated extending the ideas from Section A1.3.4

Table A8: Initialization phase definition: model policies introduced to fit the current lockdown time-series in Austria

| date | policy event | interpretation | parameters |
|---|---|---|---|
| Mar.10 | reduction of leisure time contacts | Due to the raising anxiety and awareness among the population resulting from the medial coverage, we assume that many Austrians have already minimized their contact behavior before introduction of the policies. | A simulated leisure time contact is refused by an agent with probability 0.16. |
| Mar.10 | increased hygiene and distancing in *leisure time* | Increasing awareness leads to increased hygiene among the population. | *Leisure time transmission probability* is reduced by 50%. |
| Mar.14 | closing of *schools* and *workplaces* | Although closure of schools and workplaces was announced for March 16$^{\text{th}}$, a Monday, the policy is actually active already two days earlier, as all schools and most workplaces are closed during the weekend anyway. | All *schools* and 50% of the *workplaces* are disabled for contacts. |
| Mar.14 | increased hygiene and distancing at *workplaces* | Due to increasing hygiene and distancing at work, contacts are less infectious. | *Workplace transmission probability* is reduced by 50%. |
| Mar.14 | reduced contact frequency at *workplaces* | Due to increased use of home-office, also the total number of contacts at work per day is reduced. | The average number of *workplace contacts per day* is reduced by 50%. |
| Mar.16 | reduction of leisure time contacts | Due to massive restriction of mobility, leisure time contacts of people are heavily reduced. | A simulated leisure time contact is refused by an agent with probability 0.7. |

Table A9: Lift of the lockdown measures on May 1st 2020 and introduction of the tracing strategy.

| date | policy event | interpretation | parameters |
|---|---|---|---|
| before May 01 | lockdown | All shutdown policies are active. | all policies introduced in Table A8 are active. |
| May 01 | open schools and workplaces | All policies introduced on Mar 14 are lifted. | All *school* and *workplace* locations are open for contacts as normal. Transmission probabilities and contact numbers are reset to the original value. |
| May 01 | increase of leisure tome contacts | All policies introduced on Mar 10 and Mar 16 are lifted. | *leisure time contact reduction* and *leisure time transmission probability* are fully reset to the original value. |
| May 15 | increased hygiene in *leisure time*,*schools* and *workplaces* | increase of infectivity of the disease in all settings except from households. | *leisure time/school/workplace transmission probability* are set to a calibrated value (see *stagnation levels* in Section 2.5 in the main text). |
| May 15 | start of tracing | in all strategies (except from the *no tracing* strategy) a tracing policy is introduced here | see Section 2.5 in the main text. |

### References


[1] Apache commons rng homepage. https://commons.apache.org/proper/commons-rng/. Accessed: 2020-04-17.

[2] Covid-19 information page by ages. https://www.ages.at/en/wissen-aktuell/publikationen/epidemiologische-parameter-des-covid19-ausbruchs-oesterreich-2020/. Accessed: 2020-04-08.

[3] dwh gmbh news entry for the abt simulation framework. http://www.dwh.at/en/news/the-power-of-the-abt-simulation-framework/. Accessed: 2020-04-17.

[4] Geo- and TopoJSON files of municipalities, districts and states in Austria by Flooh Perlot. Accessed: 2019-05-12.

[5] SARS-CoV-2 Steckbrief zur Coronavirus-Krankheit-2019 (COVID-19).

[6] Web-page of the City of Vienna. Acessed 2020-03-01.

[7] Statistik Austria. Betreuungs- und Pflegedienste.

[8] Statistik Austria. Bevölkerungsstand und Bevölkerungsveränderung, 2019.

[9] Statistik Austria. Bildung - Bundesanstalt Statistik Österreich, 2019.

[10] Statistik Austria. Pendlerinnen und Pendler - Bundesanstalt Statistik Österreich, 2019.

[11] Wien Statistik Austria. *Arbeitsstättenzählung 2001.* Verlag Österreich, 2004.

[12] Fabio Bellifemine, Agostino Poggi, and Giovanni Rimassa. Jade–a fipa-compliant agent framework. In *Proceedings of PAAM*, volume 99, page 33. London, 1999.

[13] Martin Bicher. *Classification of Microscopic Models with Respect to Aggregated System Behaviour.* Dissertation, TU Wien, Vienna, Austria, November 2017.

[14] Martin Bicher, Christoph Urach, and Niki Popper. GEPOC ABM: A Generic Agent-Based Population Model for Austria. In *Proceedings of the 2018 Winter Simulation Conference*, pages 2656–2667, Gothenburg, Sweden, 2018. IEEE.

[15] Martin Bicher, Matthias Wastian, Dominik Brunmeir, Matthias Rößler, and Niki Popper. Review on Monte Carlo Simulation Stopping Rules: How Many Samples Are Really Enough? In *Proceedings of the 10th EUROSIM Congress on Modelling and Simulation*, Logrono, Spain, July 2019. In Print.

[16] dwh GmbH. Technical Model Specification of the dwh and TU Wien Agent-Based Covid-19 Model. Acessed 2020-0923.

[17] Care and Consumer Protection Federal Ministry of Social Affairs, Health. Datenplattfrom COVID-19, September 2020.

[18] Care and Consumer Protection Federal Ministry of Social Affairs, Health. Rechtliche Grundlagen und Meldung übertragbarer Krankheiten, September 2020.

[19] Aneta Jadwiga Florczyk, Stefano Ferri, Vasileios Syrris, Thomas Kemper, Matina Halkia, Pierre Soille, and Martino Pesaresi. A new European settlement map from optical remotely sensed data. *IEEE Journal of Selected Topics in Applied Earth Observations and Remote Sensing*, 9(5):1978–1992, 2015. Publisher: IEEE.

[20] Aneta Jadwiga Florczyk, Stefano Ferri, Vasileios Syrris, Thomas Kemper, Matina Halkia, Pierre Soille, and Martino Pesaresi. A new european settlement map from optical remotely sensed data. *IEEE Journal of Selected Topics in Applied Earth Observations and Remote Sensing*, 9(5):1978–1992, 2016.

[21] Ilya Grigoryev. *AnyLogic 6 in three days: a quick course in simulation modeling.* AnyLogic North America, [Hampton, NJ], 2012.

[22] Volker Grimm, Uta Berger, Finn Bastiansen, Sigrunn Eliassen, Vincent Ginot, Jarl Giske, John Goss-Custard, Tamara Grand, Simone K. Heinz, Geir Huse, Andreas Huth, Jane U. Jepsen, Christian Jørgensen, Wolf M. Mooij, Birgit Müller, Guy Pe'er, Cyril Piou, Steven F. Railsback, Andrew M. Robbins, Martha M. Robbins, Eva Rossmanith, Nadja Rüger, Espen Strand, Sami Souissi, Richard A. Stillman, Rune Vabø, Ute Visser, and Donald L. DeAngelis. A standard protocol for describing individual-based and agent-based models. *Ecological Modelling*, 198(1):115–126, 2006.

[23] Volker Grimm, Uta Berger, Donald L. DeAngelis, J. Gary Polhill, Jarl Giske, and Steven F. Railsback. The ODD protocol: A review and first update. *Ecological Modelling*, 221(23):2760–2768, 2010.

[24] Joel Hellewell, Sam Abbott, Amy Gimma, Nikos I Bosse, Christopher I Jarvis, Timothy W Russell, James D Munday, Adam J Kucharski, W John Edmunds, Fiona Sun, and others. Feasibility of controlling COVID-19 outbreaks by isolation of cases and contacts. *The Lancet Global Health*, 2020. Publisher: Elsevier.

[25] Juan Ignacio Latorre Jimenez. EUROSIM 2019 Abstract Volume. In *EUROSIM 2019 Abstract Volume*. ARGESIM, 2019.

[26] Stephen A Lauer, Kyra H Grantz, Qifang Bi, Forrest K Jones, Qulu Zheng, Hannah R Meredith, Andrew S Azman, Nicholas G Reich, and Justin Lessler. The incubation period of coronavirus disease 2019 (COVID-19) from publicly reported confirmed cases: estimation and application. *Annals of internal medicine*, 2020.

[27] David Masad and Jacqueline Kazil. Mesa: an agent-based modeling framework. In *14th PYTHON in Science Conference*, pages 53–60, 2015.

[28] Makoto Matsumoto and Takuji Nishimura. Mersenne twister: a 623-dimensionally equidistributed uniform pseudo-random number generator. *ACM Transactions on Modeling and Computer Simulation (TOMACS)*, 8(1):3–30, 1998.

[29] Joël Mossong, Niel Hens, Mark Jit, Philippe Beutels, Kari Auranen, Rafael Mikolajczyk, Marco Massari, Stefania Salmaso, Gianpaolo Scalia Tomba, Jacco Wallinga, and others. POLYMOD social contact data. 2017.

[30] Muaz A. K. Niazi, Amir Hussain, and Mario Kolberg. Verification and Validation of Agent Based Simulations using the VOMAS (Virtual Overlay Multi-agent System) Approach. volume 494. CEUR-WS, July 2009.

[31] Michael J North, Thomas R Howe, Nick T Collier, and Jerry R Vos. The repast simphony runtime system. In *Proceedings of the agent 2005 conference on generative social processes, models, and mechanisms*, volume 10, pages 13–15. Citeseer, 2005.

[32] Marina Pollán, Beatriz Pérez-Gómez, Roberto Pastor-Barriuso, Jesús Oteo, Miguel A Hernán, Mayte Pérez-Olmeda, Jose L Sanmartín, Aurora Fernández-García, Israel Cruz, Nerea Fernández de Larrea, and others. Prevalence of SARS-CoV-2 in Spain (ENE-COVID): a nationwide, population-based seroepidemiological study. *The Lancet*, 396(10250):535–544, 2020. Publisher: Elsevier.

[33] S. Tisue and U. Wilensky. NetLogo: A simple environment for modelling complexity. pages 16–21, 2004.

Table A2: List of contact specific parameters (1/2). Note that all parameter values are specified for the standard model without policies. The Γ-distribution is given as $\Gamma(k, \theta)$.

| parameter | description | value | source |
|---|---|---|---|
| *leisure time contacts per day* | number of leisure time transmission-relevant contacts of an agent per day | $X \sim \Gamma(6.11, 1.0)$ | based on the results of the POLYMOD study [29] |
| *workplace contacts per day* | number of transmission-relevant contacts at work (if assigned) of an agent per day | $X \sim \Gamma(5.28, 1.0)$ | based on the results of the POLYMOD study [29] |
| *care-home contacts per day* | number of transmission-relevant contacts in a care-home (if assigned) of an agent per day | $X \sim \Gamma(5.28, 1.0)$ | based on the results of the POLYMOD study for workplaces [29] |
| *same-group probability in care-homes* | probablity to have a contact with a person of the same group (worker, inhabitant) within a care-home | 25% | Estimate |
| *school contacts per day* | number of transmission-relevant contacts at school (if assigned) of an agent per day | $X \sim \Gamma(4.64, 1.0)$ | based on the results of the POLYMOD study [29] |
| *household sizes and structure* | distribution of household sizes and structure | see source | distribution and structure from freely accessible tables for household statistics from the Austrian National Statistics Institute [8] |
| *school sizes* | The actual number of schools and pupils were gathered to calculate the average size of schools for the model. Based on this average, sizes for the simulation are sampled truncated normally. | $X \sim N(\mu, \sqrt{\mu})$, with $\mu = \frac{pupils}{schools}$ and $X > \mu/4$ a.s. | counts gathered from a publication of the Austrian National Statistics Institute [9] |
| *workplace sizes* | discrete distribution of workplace sizes | see source | gathered from a survey [11] by the Austrian National Statistics Institute |
| *care-home units* | The actual number of care-home staff and residents were gathered to calculate the number of care-home units, given a maximum capacity of 20 residents | see source | counts gathered from freely accessible tables from the Austrian National Statistics Institute [7] |
| *unemploy-ment rate* | fraction of adults who are not assigned a workplace | 10.4% | according to Austrian definition (AMS) gathered from the web-page of the city of Vienna [6] |

Table A3: List of contact specific parameters (2/2). Note that all parameter values are specified for the standard model without policies.

| parameter | description | value | source |
|---|---|---|---|
| *regional distribution of leisure time contacts* | leisure time contact partners are sampled based on origin-destination matrices on municipality level | Average fraction of all stays of persons from municipality X within municipality Y for all municipalities X and Y of Austria | gathered from mobile phone data evaluated for January 2020 |
| *regional distribution of schools* | schools are assigned based on locally biased distribution according to the regional structure: municipality ($p_m$), district($p_d$), federal-state ($p_f$) and anywhere ($p_a$) | $[p^s_m, p^s_d, p^s_f, p^s_a] = [0.66, 0.15, 0.13, 0.06]$ | data from the Austrian Bureau of Statistics [9] |
| *regional distribution of workplaces* | workplaces are assigned based on locally biased distribution according to the regional structure: municipality ($p_m$), district($p_d$), federal-state ($p_f$) and anywhere ($p_a$) | $[p^w_m, p^w_d, p^w_f, p^w_a]$ per federal state: AT1:$[0.26, 0.21, 0.16, 0.37]$ AT2:$[0.46, 0.16, 0.30, 0.08]$ AT3:$[0.29, 0.20, 0.22, 0.29]$ AT4:$[0.35, 0.24, 0.34, 0.07]$ AT5:$[0.46, 0.21, 0.26, 0.07]$ AT6:$[0.42, 0.23, 0.27, 0.08]$ AT7:$[0.41, 0.30, 0.25, 0.04]$ AT8:$[0.37, 0.35, 0.26, 0.02]$ AT9:$[0.01, 0.04, 0.84, 0.11]$ | according to commuting behaviour data from the Austrian Bureau of Statistics [10] |
| *regional distribution of care-home for residents* | care-homes are assigned based on geographical proximity to the resident | | based on expert opinions |

Table A4: List of disease specific parameters (1/3).

| parameter | description | value | source |
|---|---|---|---|
| *infection probability* | probability that a contact between a susceptible and an infected agent leads to a transmission. Depending on the type of contact (household, school, work, leisure time), this parameter is scaled by a factor | $\alpha_{\text{household}} = 0.25$, $\alpha_{\text{workplace}} = \alpha_{\text{school}} = \alpha_{\text{leisure time}} = \alpha_{\text{care-home}} = 0.05$ | calibrated based on the doubling rates in Austria before introduction of policies (see Section Calibration in [16]). |
| *incubation time* | time between infection and symptom on-set | scaled $\beta$ distribution with $min(X) = 2[d]$, $\max(X) = 14[d]$, $E(X) = 5.1[d]$ | based on [26] |
| *latency time* | time between infection and infectivity | always incubation time minus $2[d]$ | estimates by the Robert Koch Institute ( [5], Section 8) |
| *reaction time* | time between symptom on-set and testing of the agent which furthermore leads to its confirmation and home isolation | $X \sim \text{Weibull}(4.29, 1.65)$ | based on [24] |
| *infectious duration* | duration agents are infectious | $X \sim \text{Tri}(3, 6, 9)$ | expert estimates (Austrian virologists) |
| *hospitalisation delay* | time between start of home isolation and hospitalisation | $X \sim \text{Tri}(0, 0.5, 1.5)$ | expert opinions (hospital administrators) |
| *ICU delay* | time between start hospitalisation and transfer to ICU | $X \sim \text{Tri}(0, 0.5, 1.5)$ | expert opinions (hospital administrators) |

Table A5: List of disease specific parameters (2/3).

| parameter | description | value | source |
|---|---|---|---|
| *recovery time home quarantined* | time between symptom on-set and recovery for symptomatic persons in home isolation | $X \sim Tri(11, 14, 16)$ | based on expert opinions and calibrated w.r.t. official recovery data |
| *recovery time severe* | time between symptom on-set and recovery for symptomatic persons in normal hospital beds | $X \sim$ Weibull$(20.0, 1.8)$ | Cox regression on reported data by Austrian hospitals (Epidemiologisches Meldesystem [17]) |
| *recovery time critical* | time between symptom on-set and recovery for symptomatic persons in ICU beds | $X \sim$ Weibull$(35.0, 2.0)$ | Cox regression on reported data by Austrian hospitals (Epidemiologisches Meldesystem [17]) |
| *recovery time unconfirmed* | time between symptom on-set and recovery for unconfirmed persons (usually asymptomatic) | $Tri(1, 5, 7)$ | based on expert opinions |
| *detection probability* | probability of an infected person to get detected by a test | [ 3.0, 9.0, 22.0, 20.0, 24.0, 28.0, 20.0, 21.0, 33.0, 58.0 ]% for 10 year age-classes | Age distribution is gathered by comparison of Austria's age pyramid with the distribution of the confirmed cases. The overall detection probability is calibrated to 19.5% based on a recent antibody study from Spain [32]. |
| *hospitalisation probability* | age-dependent probability that a detected patient requires hospitalisation | 2020, Mar-Jul: [ 0.0, 4.8, 1.1, 3.3, 4.5, 8.8, 33.1, 33.1, 58.1, 71.4, 62.2 ]%, from Aug 2020 : [ 0.0, 4.6, 2.7, 5.2, 5.2, 6.8, 10.5, 16.9, 32.6, 50.0, 40.8 ]% for 10 year age-classes | Distribution is based on comparison of the age distribution of confirmed cases with the age distribution of hospitalised cases (Epidemiologisches Meldesystem [17]). |

Table A6: List of disease specific parameters (3/3).

| parameter | description | value | source |
|---|---|---|---|
| *icu probability* | probability that a hospitalised agent becomes critical (needs an intensive care unit) | 16.6% | calibrated for Austrian ICUs with data from the ministry of internal affairs |
| *death-by-COVID-19 probability - unconfirmed* | probability that an unconfirmed infected agent dies due to COVID-19 | 0% | No data is available. Moreover, unconfirmed COVID-19 cases are typically asymptomatic. |
| *death-by-COVID-19 probability - mild* | probability that a non-hospitalized (mild) infected agent dies due to COVID-19 | 2020, Mar-Jul: [ 0.0, 0.0, 0.0, 0.0, 0.1, 0.2, 1.6, 7.5, 20.3, 26.7 ]%, from Aug 2020 : [ 0.0, 0.0, 0.0, 0.0, 0.0, 0.1, 0.2, 1.2, 4.5, 9.0 ]%, for 10 year age-classes | Distribution is based on comparison of the age distribution of confirmed non-severe and non-critical cases with the age distribution of fatal cases (Epidemiologisches Meldesystem [17]). |
| *death-by-COVID-19 probability - severe* | probability that an infected agent requiring a normal bed (severe) dies due to COVID-19 | 2020, Mar-Jul: [ 0.0, 0.0, 0.0, 0.0, 0.9, 3.3, 8.3, 24.3, 31.5, 43.5 ]% , from Aug 2020 : [ 0.0, 0.0, 0.0, 0.0, 0.0, 0.0, 1.2, 5.1, 11.4, 14.3 ]% ,for 10 year age-classes | Distribution is based on comparison of the age distribution of confirmed severe cases with the age distribution of fatal cases (Epidemiologisches Meldesystem by AGES [17]). |
| *death-by-COVID-19 probability - critical* | probability that an infected agent requiring an ICU bed (critical) dies due to COVID-19 | 2020, Mar-Jul: [ 0.0, 0.0, 0.0, 0.0, 6.3, 18.8, 26.7, 34.6, 63.3, 66.7 ]%, from Aug 2020 : [ 0.0, 0.0, 0.0, 0.0, 1.7, 0.0, 6.8, 12.1, 11.1, 28.6 ]%, for 10 year age-classes | Distribution is based on comparison of the age distribution of confirmed critical cases with the age distribution of fatal cases (Epidemiologisches Meldesystem [18]). |

Table A7: List of possible event-timeline elements that can pose for the model's input including their effect and, if available, options for the event parametrisation.

| event | description | parameters |
|---|---|---|
| *leisure-time contact number reduction event* | Based on an age-class (child, adult, retired) dependent probability, an agent may "reject" a leisure-time contact with a different agent. As the rejection happens symmetrically, the probabilities multiply. | age-class-dependent fraction by which daily leisure-time contacts are reduced |
| *location closing event* | Fraction of locations of a certain type are closed in this policy. | affected location type; fraction of locations of this type that remain open / are opened |
| *adapt hygiene event* | Changes the *infection probability* for contacts in a certain location | affected location type; factor to scale the original *infection probability* with |
| *start location tracing event* | Starts with location tracing measures. I.e. all members of a newly *confirmed* agent's location are put under preventive isolation for a certain period of time. | affected location type; length of preventive quarantine length |
| *start contact tracing event* | Starts with contact tracing measures. I.e. recorded contacts of a newly *confirmed* agent are put under preventive isolation. | fraction of agents capable of recording contacts; length of preventive quarantine length |
